# Utility of glucose, lipid and kidney function Trajectory Measures for incident Cardiovascular Disease risk prediction for people living with Type 2 Diabetes: a case-study using Danish registry data

**DOI:** 10.64898/2026.03.06.26347493

**Authors:** Peter P. Harms, Omar Silverman-Retana, Jonas Schaarup, Marieke T. Blom, Anders A. Isaksen, Daniel R. Witte

## Abstract

**Introduction:** Cardiovascular disease (CVD) is an important complication of type 2 diabetes (T2D). Current incident CVD-prediction models use single baseline measurements and achieve moderate performance in people with T2D, with C-indices around 0.7. Modern healthcare registries contain repeated measurements of HbA1c, LDL-cholesterol and eGFR, which could carry incremental predictive value. However, the added value of trajectory measures for CVD-risk prediction remains unclear. We aimed to investigate the utility of HbA1c, LDL-cholesterol and eGFR trajectory measures for incident CVD-risk prediction in people with T2D.

**Methods:** We studied 83,326 people with T2D from Danish nation-wide registers, who were without a CVD-history at baseline (January 1st 2015), and had ≥2 recorded HbA1c, LDL-cholesterol and eGFR measurements between 2012-2014. Their last measurement was considered as baseline. Across 2012-2014, three types of paired trajectory measures were calculated for each participant (mean & standard deviation (SD), median & interquartile range (IQR), and intercept & slope from a fitted growth model), for HbA1c, LDL-cholesterol, and eGFR, respectively. Reference Cox-regression models for CVD-events (ICD-10 codes assessed prospectively from 2015- 2020) included only baseline measurements (age, sex , age at T2D onset, HbA1c, LDL-cholesterol, HDL-cholesterol, eGFR, and medication use). Next, the paired trajectory measures were sequentially added to the reference model, computing Hazard Ratios, C-indices and Net reclassification index (NRI) with 95% confidence intervals. Lastly, a combined model was fitted.

**Results:** At baseline, mean age was 65 (SD±12), median HbA1c was 48 (mmol/mol, IQR43-56), and 48% were female. During a median 6 years of follow-up 11,280 (14%) people had a CVD-event (ischemic heart disease: 40%; stroke: 32%; heart failure: 24%; CVD-mortality: 5%). Accounting for the reference model, trajectory measures of dispersion and change were associated with CVD-events, with hazard ratios ≈ 1.1 for HbA1c and eGFR, and >1.4 for LDL-cholesterol. Measures centrality did not show an association with CVD events.

Addition of trajectory measures produced minimal gains in discrimination (C-index Δ +0.001-+0.003) but modest improvements in net reclassification (continuous NRI ≈ +3- +9%).

**Conclusions:** Trajectory dispersion or change measures for HbA1c, eGFR and especially LDL-cholesterol, easily obtained from routine data, might moderately enhance incident CVD-risk prediction in people with T2D.

## Introduction

Cardiovascular disease (CVD) is an important and frequent complication of type 2 diabetes (T2D), affecting approximately one in three persons living with the disease. People living with T2D have a roughly twofold increased risk of CVD compared with individuals without T2D.^1^ Cardiovascular complications contribute to 30-50% of all mortality in people living with T2D.^2,3^ Nonetheless, not all people living with T2D have equal CVD-risk as the degree of adequate glycemic control and other CVD-risk factors vary considerably within each person.^4^

Individual stratification based on CVD-risk prediction models^5,6^ is a central component of current cardiometabolic treatment strategies. These models are used by clinicians not only to guide decisions but also to inform people living with T2D about their risk and the impact of treatment. Currently, these prognostic prediction models use single measurements of risk factors at one single time-point (i.e. the most recent value: baseline), and they show only moderate performance in people living with T2D, with C-indices around 0.7 in validation studies (theoretical range: 0.5–1.0).^7,8^ Moreover, the models also tend to underestimate CVD-risk in certain groups, specifically in women and young people.^9,10^

These limitations underscore the need for improving CVD-risk prediction models to more accurately identify individuals living with T2D at high risk of developing CVD. Repeated measurements may provide incremental predictive value beyond single baseline measurements.^11^ Single measurements can yield inaccurate risk prediction estimates, when a risk factor is measured with substantial error or has high within-person variability. Additionally, integrating information on the rate of change or variability in a risk factor over time might improve CVD-risk prediction overall and for specific age and sex subgroups. By utilizing longitudinal data contained in healthcare registries we might get more out of the data that is already readily available.

Indeed, a few cohort studies highlight the incremental predictive value of using trajectory measures derived from repeated measurements over time for incident CVD-risk prediction in the general population. A meta-analysis of the Emerging Risk Factors Collaboration studies (191,445 persons aged 40-79) demonstrated that using trajectory measures of systolic blood pressure, total cholesterol and HDL cholesterol noticeably improved risk discrimination and (re)classification, compared to single baseline measurement models.^12^ The Atherosclerosis Risk in Communities (ARIC) study observed similar improvements in discrimination with trajectory measures of systolic blood pressure.^13^ Lastly, the Korean National Health Screening Cohort (467,708 persons aged 40-70) even reported substantial improvement in discrimination and especially (re)classification after incorporating trajectory measures for body mass index, systolic blood pressure, diastolic blood pressure, total cholesterol, fasting plasma glucose, smoking, and exercise.^14^

However, to date no studies have investigated the utility of trajectory measures for incident CVD-risk prediction specifically in people living with T2D, none have examined trajectory measures of key risk factors such as glycated hemoglobin (HbA1c), low-density lipoprotein cholesterol (LDL-C), and estimated glomerular filtration rate (eGFR), and none have used modern healthcare registries that offer a wealth of readily available longitudinal data, including repeated measurements of HbA1c, LDL-cholesterol and eGFR, coupled with population-wide prospective assessment of CVD outcomes. Furthermore, the impact of trajectory measures on predictive performance in distinct sex and age groups remains underexplored.

Therefore, we aimed to investigate the utility of adding HbA1c, LDL-C and eGFR trajectory measures to incident CVD-risk prediction models for people with T2D using routinely collected data from nationwide registries.

## Methods

### Study design and data sources

We conducted a register-based multivariable prognostic prediction modeling study which comprised development and internal validation (using resampling) of CVD-risk prediction models updated with trajectory measure predictors.^15^ Data from Danish nationwide registries were linked using the unique personal identifier given to all Danish residents at birth or immigration.^16^ The Danish public healthcare system is tax funded and offers universal access to tax-paid medical services from hospitals, specialists, and general practitioners for all residents, free of cost at the point of care.^17^ The clinical and population registers thus provide nearly complete population-wide follow-up for incident events and mortality.

Information on age, sex and vital status was obtained from the Danish Civil Registration System.^16^ Information on primary and secondary diagnoses recorded during hospital admissions and outpatient contacts was obtained from the Danish National Patient Register, classified according to International Classification of Diseases, 8^th^ revision (ICD-8, from 1977 until 1994) and 10^th^ revision (ICD-10, since 1994).^18^ Information on routine clinical laboratory test results was obtained from the Register of Laboratory Results for Research (RLRR), classified according to the Nomenclature for Properties and Units (NPU) terminology. From 2012-2015 the RLRR covered four of five Danish administrative regions (excluding the Southern Denmark region). Information on medication use was obtained from the Danish National Prescription Registry, classified according to the Anatomical Therapeutic Chemical (ATC) classification since 1995.^19^ Information on specific causes of death were obtained from the Danish National Death Registers, classified according to the ICD-10 since 2002. A detailed description of all codes used in this study to define variables is available in supplementary Table S1.

### Participants and study sample

The source population comprised all 5,707,176 Danish residents recorded in the Danish Civil Registration System as alive on the 1^st^ of January 2015, which was chosen as the baseline index date. Next, we selected the 229,759 persons with prevalent T2D at baseline, using the previously validated Open-Source Diabetes Classifier.^20^ This algorithm identifies T2D based on HbA1c measurements ≥48 mmol/mol (6.5%), hospital diagnoses of diabetes, diabetes-specific podiatrist services, and purchases of glucose lowering medication (excluding semaglutide, dapagliflozin and empagliflozin). The second date of these occurrences is taken as diabetes incidence date. Type 1 diabetes and T2D are distinguished based on hospital diagnoses of diabetes and patterns of insulin purchases.

We further excluded 76,180 (33.1%) persons without ≥2 records of HbA1c, cholesterol and eGFR measurements within the 3 years prior to the baseline index date (2012, 2013 and 2014). This included 32,432 (42.6%) persons from the Southern Denmark region that was not covered by the RLRR laboratory register during this period. The final study sample for analyses included 83,326 individuals (Figure 1).

**Figure 1.**
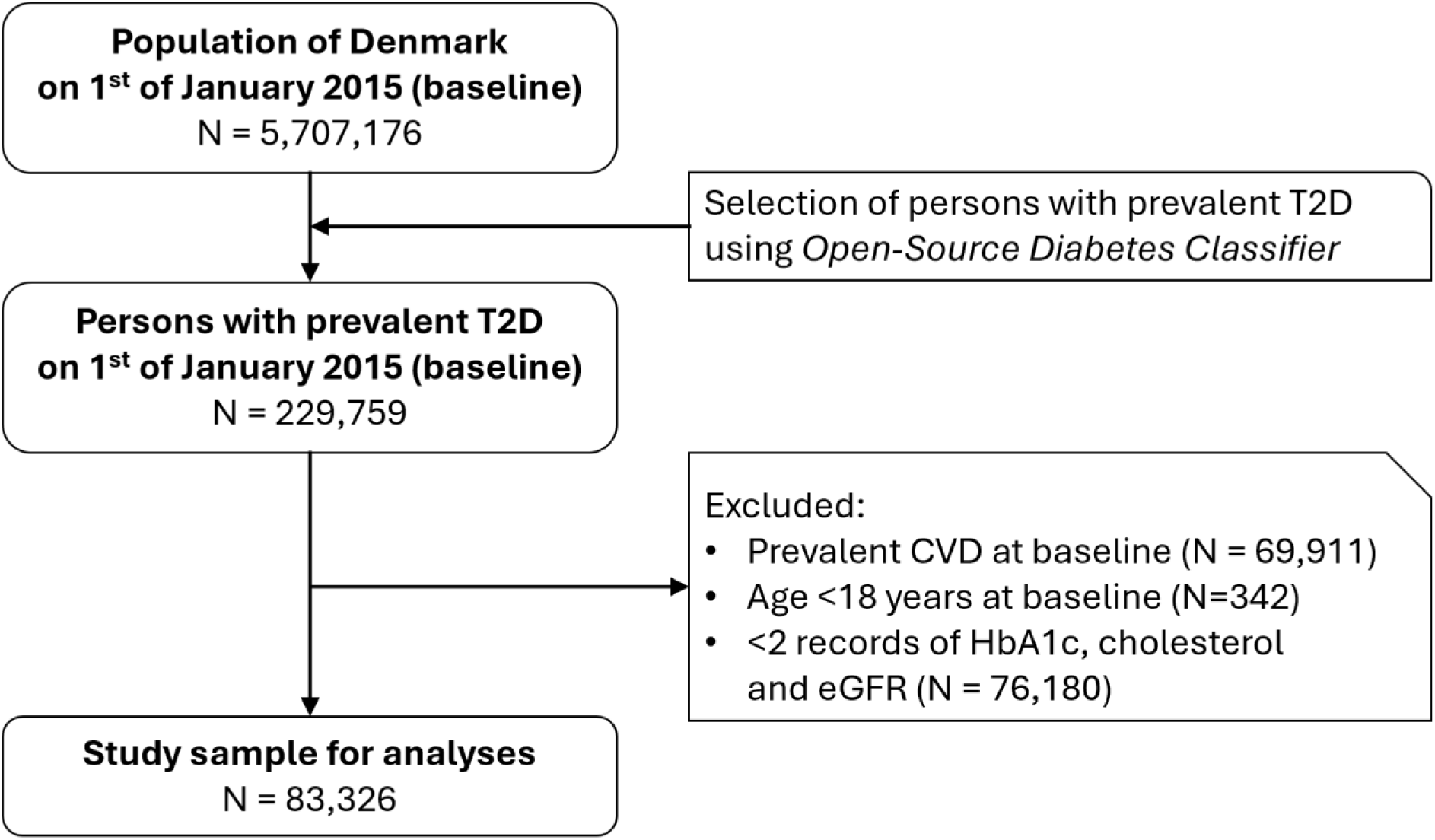
Flowchart of study sample derivation.

### Baseline predictors

Age at baseline (years), age at T2D diagnosis (years) and sex (female/male) were defined as continuous and binary variables, respectively. HbA1c (mmol/mol), LDL-cholesterol (mmol/L), HDL-cholesterol (mmol/L) and eGFR (mL/min/1.73m^2^) levels at baseline were considered the most recent measurement before the baseline index date (1^st^ of January 2015) and defined as continuous variables. Blood pressure lowering, lipid lowering, oral glucose lowering, and insulin medication use at baseline were defined as at least one purchase during the 12 months before the baseline index date. These four types of medication use were all defined as binary variables (no/yes).

### Trajectory measure predictors

Three types of paired trajectory measures were calculated for each individual participant across all HbA1c, LDL-cholesterol and eGFR measurements within the 3 years prior to the baseline index date (2012, 2013 and 2014), respectively. These trajectory measures included individual mean & SD, individual median & IQR, and individual intercept & slope from a fitted growth model. The growth models were fitted using a linear mixed/multi-level model with a random intercept and a random slope on the participant level, with time (years) before baseline as the independent variable and HbA1c, LDL-cholesterol or eGFR as the dependent variable, respectively. The intercept represents the estimated individual HbA1c, LDL-cholesterol or eGFR level at baseline index date, and the slope is interpreted as the estimated individual average annual change in the respective variable going forward in time up to baseline.

### Cardiovascular outcome

Non-fatal CVD-events after baseline were identified through the Danish National Patient Register and defined using primary and secondary diagnosis codes (ICD-10) for ischemic heart disease (including angina pectoris and myocardial infarction), stroke, or heart failure (and associated surgical intervention procedure codes). Cause of death was determined using the Danish National Death Register based on the underlying cause of death diagnosis codes (ICD-10), and classified as CVD-mortality (ischemic heart disease, stroke, or heart failure) or non-CVD death.

The endpoint of Major Adverse Cardiovascular Events (MACE) comprised non-fatal events of ischemic heart disease, stroke, heart failure, and CVD-mortality. Follow-up time was defined as the period from baseline until the first MACE, non-CVD death or end of study (31th December 2021), whichever occurred first.

### Statistical analysis

All analyses were complete-case analyses. Baseline characteristics were summarized in means, medians, or proportions, as appropriate. Summary statistics for the trajectory measures, along with the number of repeated measurements and their timespan were calculated in means, medians, frequencies or days, as appropriate. Follow-up time and incidence of MACE was calculated and graphically presented using cumulative incidence curves. All analyses were performed for the total study sample and in additional exploratory analyses stratified by age group (<40/40-60/>60), sex and blood pressure lowering, lipid lowering, oral glucose lowering medication, and insulin use.

Reference Cox-regression models were fitted for the MACE endpoint in the total study sample and stratified by age group, sex and medication use, respectively. All reference models included baseline measurements for age, age at T2D diagnosis, sex, HbA1c, LDL-cholesterol, HDL-cholesterol, eGFR, blood pressure lowering, lipid lowering, oral glucose lowering medication and insulin use.

Next, the trajectory-measures-models were fitted using an offset procedure, sequentially adding one of the three types of paired trajectory measures to the respective (sub)sample reference model: mean & SD, median & IQR, and intercept & slope. Hazard ratios (HRs) with 95% confidence intervals (95%CI) were computed for the pairs of trajectory measures from all the trajectory-measures-models, to assess the association of trajectory measures with the endpoint adjusted for all baseline predictors in the reference model.

Model discriminative performance and its improvement were assessed using the C-index and continuous NRI. The change in C-index was obtained by subtracting the calculated C-index of the reference model from the C-index of the corresponding trajectory-measures-model. The continuous NRI was determined by summing the increases in predicted risk to any extent for people with the endpoint under the model with trajectory measures compared with the corresponding reference model, and similarly, the risk decreases for people without the endpoint. Bootstrapping was used to calculate 95%CIs around C-index and continuous NRI.

Finally, to assess the combined effect of trajectory-measures-predictors on model performance in the total sample, we constructed a model incorporating three sets of paired trajectory measures for HbA1c, LDL-cholesterol, and eGFR each. Selection of the sets of paired trajectory measures (mean & SD, median & IQR or intercept & slope) included in this final combined-trajectory-measures-model was based on highest NRI of the simple paired trajectory-measures-models.

Analyses were performed with R (Studio) version 4.4.1 (R Foundation for Statistical Computing, Vienna, Austria) in combination with the R packages arrow (16.1.0), tidyverse (2.0.0), gtsummary (1.7.2), survival (3.7-0), CsChange (0.1.7) and nricens (1.6)

## Results

### Baseline characteristics

At baseline, mean (±SD) age was 65 (±12) years, median (IQR) T2D duration was 6 (3-10) years, 48% were female, the majority used blood pressure lowering (76%), lipid lowering (69%) and oral glucose lowering (80%) medication, 18% used insulin, and median (IQR) HbA1c, LDL-cholesterol and eGFR were 48 (43-56) mmol/mol, 4.2 (3.6-4.9) mmol/L, and 83 (68-90) mL/min/1.73m2, respectively. (Table 1). In the additional and exploratory stratified analysis, HbA1c was higher in younger age-groups, men and those using lipid or oral glucose lowering medication and insulin. LDL-cholesterol was higher in younger age-groups and women, and lower in those using blood pressure, lipid, oral glucose lowering medication and insulin. eGFR was higher in younger age-groups, men and in those not using blood pressure or lipid lowering medication and insulin (Table S2).

**Table 1.** Baseline characteristics of study sample.

| Characteristic | Total sample |
| --- | --- |
| Number | 83,326 |
| Age (years) | 65 (±12) |
| Age at T2D diagnosis (years) | 57 (±13) |
| T2D duration (years) | 6 (3-10) |
| Female (yes) | 48% |
| HbA1C (mmol/mol) | 48 (43-56) |
| Total cholesterol (mmol/L) | 4.2 (3.6-4.9) |
| HDL-cholesterol (mmol/L) | 1.3 (1.0-1.5) |
| LDL-cholesterol (mmol/L) | 2.1 (1.6-2.8) |
| eGFR (mL/min/1.73m<sup>2</sup>) | 83 (68-90) |
| Blood pressure lowering medication use (yes) | 76% |
| Lipid lowering medication use (yes) | 69% |
| Oral glucose lowering medication use (yes) | 80% |
| Insulin medication use (yes) | 18% |
| Data are presented as mean (±SD), median (Q1-Q3), or proportion (%). Baseline measurements were defined as the most recent measurement before the baseline index date of 1<sup>st</sup> of January 2015. T2D, type 2 diabetes; HbA1c, glycated hemoglobin A1c; LDL, low-density lipoprotein; HDL, high-density lipoprotein; eGFR, estimated glomerular filtration rate. | |

### Trajectory measures characteristics

Across the three years prior to the baseline index date (2012, 2013 and 2014), median (IQR) number of measurements was 6 (4-9) for HbA1c, 3 (3-5) for LDL-cholesterol, and 6 (3-9) for eGFR, corresponding to median (IQR) observation windows of 820 (473-966), 406 (362-862) and 798 (522-956) days, respectively (Table 2). Central tendencies of individual trajectory measures (means, medians, or model intercepts) clustered narrowly around 48–49 mmol/mol for HbA1c, 2.2–2.4 mmol/L for LDL-cholesterol, and 82–84 mL/min/1.73 m² for eGFR. Dispersion of means, medians, or model intercepts was comparable across trajectory measures within each risk factor domain (HbA1c, LDL-cholesterol, and eGFR). Dispersion metrics (SD and IQR) were similar within risk factor domains, while estimated slopes were smaller in magnitude than SD and IQR, reflecting the modeled rate of within-person change over a year rather than within-person variability in repeated measurements over three years. Differences in trajectory measure summaries stratified by age group (<40/40-60/>60), sex, and medication use were consistent with patterns observed in baseline characteristics (Table S3).

**Table 2.** Summary statistics for the individual trajectory measures, along with the number of repeated measurements and their time-span.

| **Trajectory measure** | **Total sample** |
| --- | --- |
| Number | 83,326 |
| HbA1c (mmol/mol) |  |
| Number of measurements | 6 (4-9) |
| Time-span (days) | 820 (473-966) |
| mean | 49 (44-58) |
| SD | 3 (2-7) |
| median | 49 (44-56) |
| IQR | 4 (2-7) |
| intercept | 48 (44-56) |
| slope | 1 (±4) |
| LDL-cholesterol (mmol/L) |  |
| Number of measurements | 3 (3-5) |
| Time-span (days) | 706 (362-862) |
| mean | 2 (±1) |
| SD | 0.3 (0.2-0.6) |
| median | 2 (±1) |
| IQR | 0.3 (0.2-0.5) |
| intercept | 2 (±1) |
| slope | 0.1 (±0.1) |
| eGFR (mL/min/1.73m<sup>2</sup>) |  |
| Number of measurements | 6 (3-9) |
| Time-span (days) | 798 (522-956) |
| mean | 84 (70-90) |
| SD | 6 (3-9) |
| median | 84 (70-90) |
| IQR | 5 (2-9) |
| intercept | 82 (68-89) |
| slope | 1 ($\pm 3$) |

Data are presented as mean ( ± SD), median (Q1-Q3), frequencies (n) or time (days). The trajectory measures were calculated including all recorded measurements within the 3 years prior to the baseline index date of 1<sup>st</sup> of January 2015. (2012, 2013 and 2014).
Note: the trajectory measures were paired as mean & SD, median & IQR and intercept & slope from a fitted growth model.
T2D: type 2 diabetes; HbA1c: glycated hemoglobin A1c; LDL: low-density lipoprotein; eGFR: estimated glomerular filtration rate; SD: Standard deviation, IQR: Interquartile range.

### Follow-up and cardiovascular events

During a median (IQR) 6 (6-6) years of follow-up 11,280 (13.5%) people had a CVD-event (ischemic heart disease: 40.0%; stroke: 31.5%; heart failure: 23.5%; CVD-mortality: 5.0%). Incidence rates (95%CI) per 1000 person-years for the MACE endpoint were 25.5 (25.0-25.9) in the total sample, 4.4 (3.4-5.6) in the age-group below 40, 15.0 (14.3-15.6) in the 40- 60 age-group, 31.6 (31.0-32.3) in the over 60 age-group, 21.5 (20.9-22.1) for women and 29.2 (28.5-30.0) for men, while rates in the medication use strata ranged from 16.8 (16.1-17.6) in people not using blood lowering medication to 34.4 (33.0-35.7) in people using insulin (Table S4). Cumulative incidence curves of the MACE endpoint show proportional hazards over time and a similar pattern to the incidence rates (Figure S1).

### Associations with CVD-events

Accounting for the reference models’ baseline variables, trajectory measures of dispersion such as SD, IQR and slope were associated with MACE, but not trajectory measures of centrality such as mean, median and intercept (Table 3). For HbA1c, steeper slopes were consistently associated with higher risk of MACE across most subgroups, with an adjusted HR of 1.16 (1.10-1.23). Similarly, higher HbA1c SD and IQR showed important associations with higher risk of MACE, adjusted HRs of 1.08 (1.03-1.12) and 1.05 (1.02-1.08), respectively. LDL-cholesterol slope also demonstrated a strong association, with HR reaching 1.48 (1.25-1.76). Measures of LDL-cholesterol variability SD and IQR were likewise robustly associated with MACE, with HRs of 1.15 (1.08-1.21) and 1.12 (1.06-1.17), respectively. For eGFR, dispersion measures SD and IQR were also associated with MACE, with HRs of 1.20 (1.16-1.25) and 1.12 (1.09-1.15), respectively, while slope showed modest association with a HR of 1.10 (1.03-1.17). Similar patterns in HRs were observed in the additional and exploratory stratified analysis (Table S5). Details of the reference models’ coefficients and HRs are provided in supplementary material (Table S6).

**Table 3.** Association of paired trajectory measures with MACE endpoint, after adjustment for baseline predictors.

| Trajectory measure | Adjusted HR (95%CI) for MACE |
| --- | --- |
|  | Total sample |
| HbA1c<sup>1</sup> |  |
| mean | 1.01 (0.99-1.03) |
| SD | 1.08 (1.03-1.12) |
| median | 1.01 (1.00-1.03) |
| IQR | 1.05 (1.02-1.08) |
| intercept | 1.02 (1.01-1.04) |
| slope | 1.16 (1.10-1.23) |
| LDL-cholesterol<sup>1</sup> |  |
| mean | 1.01 (0.99-1.03) |
| SD | 1.15 (1.08-1.21) |
| median | 1.01 (0.99-1.04) |
| IQR | 1.12 (1.06-1.17) |
| intercept | 1.04 (1.01-1.07) |
| slope | 1.48 (1.25-1.76) |
| eGFR<sup>1</sup> |  |
| mean | 0.99 (0.98-1.00) |
| SD | 1.20 (1.16-1.25) |
| median | 1.00 (0.99-1.01) |
| IQR | 1.12 (1.09-1.15) |
| intercept | 1.00 (0.99-1.01) |
| slope | 1.10 (1.03-1.17) |

<sup>1</sup> HbA1c: per 10 mmol/mol, LDL-cholesterol: per 1 mmol/l, eGFR: per 10 ml/min/1.73m<sup>2</sup>.
Data are presented as HR (95%CI) for MACE: Ischemic heart disease, stroke, heart failure or CVD-mortality.
Note: The trajectory measures were calculated including all recorded measurements within the 3 years prior to the baseline index date of 1<sup>st</sup> of January 2015. (2012, 2013 and 2014) and paired as mean & SD, median & IQR, and intercept & slope from a fitted growth model. The trajectory measure models were fitted using an offset procedure, sequentially adding one of the three types of paired trajectory measures to the respective (sub)sample reference model: mean & SD, median & IQR, and intercept & slope. All reference models (Cox-regression) included baseline measurements for age, age at T2D diagnosis, sex, HbA1c, LDL-cholesterol, HDL-cholesterol, eGFR, blood pressure lowering, lipid lowering, oral glucose lowering medication and insulin use.
HbA1c: Hemoglobin A1c, LDL: low density lipoprotein, eGFR: estimated glomerular filtration rate, HR: Hazard ratio, CI: Confidence Interval, SD: Standard deviation, IQR: Interquartile range.

### Prediction model performance

The reference model showed moderate diagnostic accuracy with a C-index of 0.67 (0.66-0.67) for the total sample. C-index change showed none or only minimal improvements in the range of +0.001-0.002 across the trajectory measures updated models (Table 4). The continuous NRI consistently showed modest risk classification improvement in the total sample across all pairs of trajectory measures except one with NRIs ranging from +3.3% (0.2-7.2) for the LDL-cholesterol Median & IQR updated model to +7.9% (4.6-11.5) for the HbA1c Intercept & slope model. Only the model updated with intercept & slope of eGFR which did not show a substantial improvement in NRI (+3.1 %(-3.0-8.5)). To illustrate these NRIs in a practical example: if a model including LDL-cholesterol Median & IQR was used in 1.000 T2D patients, a net 33 more patients would be correctly (re)classified as being at higher risk (or lower as appropriate) compared to the reference model. Likewise using a model including HbA1c Intercept & slope would correctly (re)classify 79 additional patients.

**Table 4.**
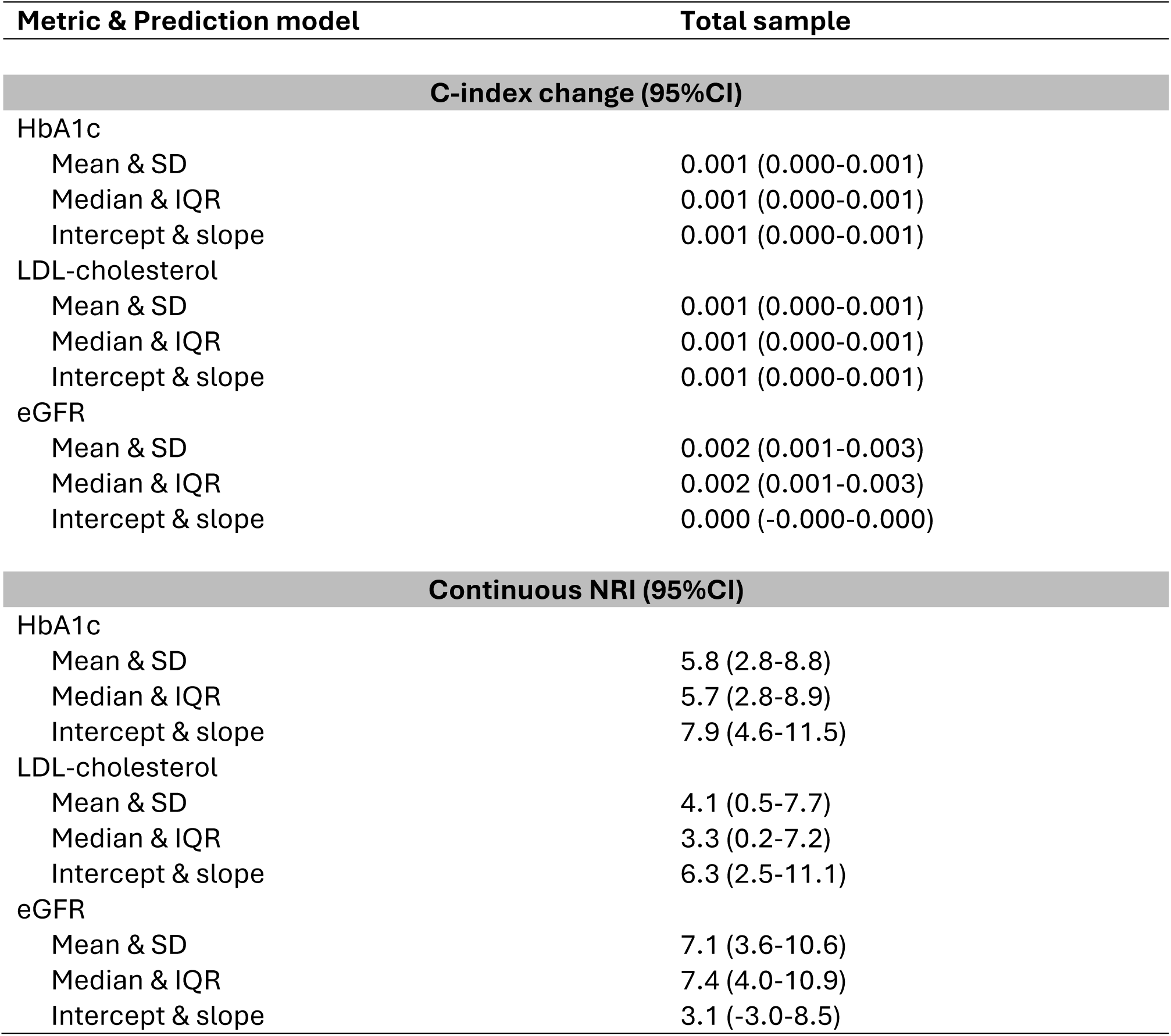
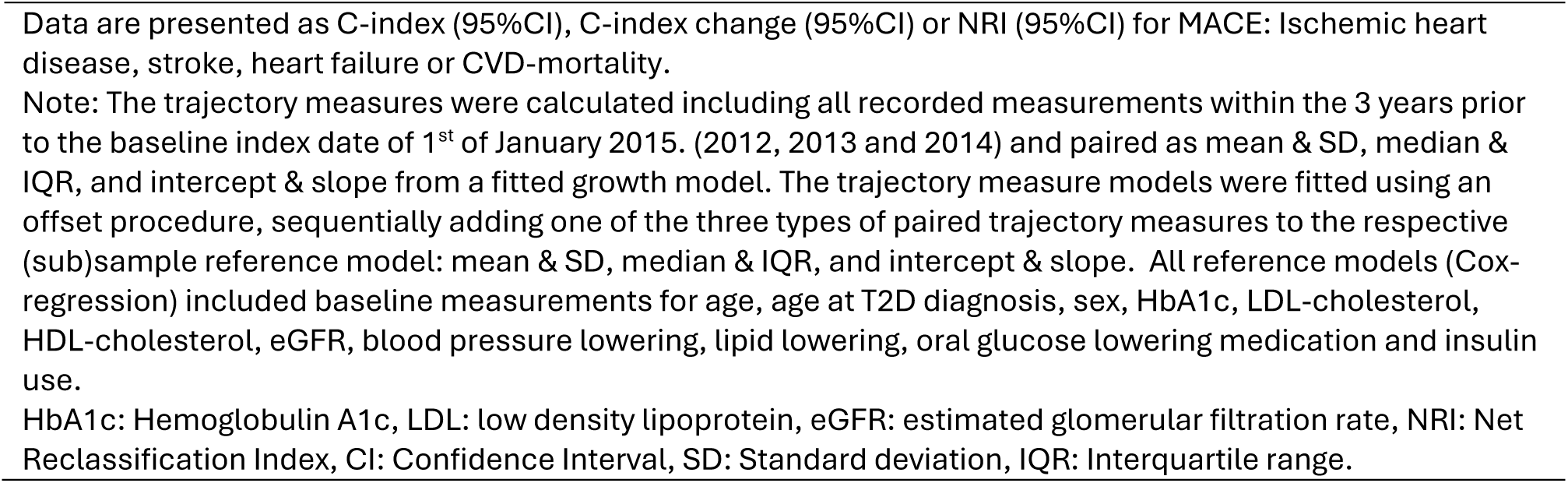
Performance and reclassification statistics of reference and trajectory measure updated prediction models.

| Metric & Prediction model |  | Total sample |
| --- | --- | --- |
| **C-index change (95%CI)** |  |  |
| HbA1c |  |  |
| Mean & SD |  | 0.001 (0.000-0.001) |
| Median & IQR |  | 0.001 (0.000-0.001) |
| Intercept & slope |  | 0.001 (0.000-0.001) |
| LDL-cholesterol |  |  |
| Mean & SD |  | 0.001 (0.000-0.001) |
| Median & IQR |  | 0.001 (0.000-0.001) |
| Intercept & slope |  | 0.001 (0.000-0.001) |
| eGFR |  |  |
| Mean & SD |  | 0.002 (0.001-0.003) |
| Median & IQR |  | 0.002 (0.001-0.003) |
| Intercept & slope |  | 0.000 (-0.000-0.000) |
| **Continuous NRI (95%CI)** |  |  |
| HbA1c |  |  |
| Mean & SD |  | 5.8 (2.8-8.8) |
| Median & IQR |  | 5.7 (2.8-8.9) |
| Intercept & slope |  | 7.9 (4.6-11.5) |
| LDL-cholesterol |  |  |
| Mean & SD |  | 4.1 (0.5-7.7) |
| Median & IQR |  | 3.3 (0.2-7.2) |
| Intercept & slope |  | 6.3 (2.5-11.1) |
| eGFR |  |  |
| Mean & SD |  | 7.1 (3.6-10.6) |
| Median & IQR |  | 7.4 (4.0-10.9) |
| Intercept & slope |  | 3.1 (-3.0-8.5) |

Data are presented as C-index (95%CI), C-index change (95%CI) or NRI (95%CI) for MACE: Ischemic heart disease, stroke, heart failure or CVD-mortality.
Note: The trajectory measures were calculated including all recorded measurements within the 3 years prior to the baseline index date of 1<sup>st</sup> of January 2015. (2012, 2013 and 2014) and paired as mean & SD, median & IQR, and intercept & slope from a fitted growth model. The trajectory measure models were fitted using an offset procedure, sequentially adding one of the three types of paired trajectory measures to the respective (sub)sample reference model: mean & SD, median & IQR, and intercept & slope. All reference models (Cox-regression) included baseline measurements for age, age at T2D diagnosis, sex, HbA1c, LDL-cholesterol, HDL-cholesterol, eGFR, blood pressure lowering, lipid lowering, oral glucose lowering medication and insulin use.
HbA1c: Hemoglobulin A1c, LDL: low density lipoprotein, eGFR: estimated glomerular filtration rate, NRI: Net Reclassification Index, CI: Confidence Interval, SD: Standard deviation, IQR: Interquartile range.

The additional and exploratory stratified analyses suggest improvements in risk (re)classification might be more pronounced in age groups >40 years, males, and users of blood pressure, lipid, or oral glucose lowering medications but did not provide irrefutable additional insights (Table S7 & S8).

Finally, the combined trajectory measures updated model included HbA1c intercept & slope, LDL-cholesterol intercept & slope, and eGFR median & IQR. This model had a C-index of 0.67 (0.66-0.67), an improvement of 0.003 (0.002-0.004) compared to the reference model, and its NRI was 9.0% (5.0-12.9).

## Discussion

### Main findings

In this study, trajectory measures of dispersion or change (SD, IQR, slope) derived from repeated clinical measurements of HbA1c, LDL-cholesterol and eGFR were independently associated with incident MACE after adjustment for baseline risk factors, but centrality measures (mean, median, intercept) were not. Effect sizes were modest for HbA1c and eGFR (HRs ≈ 1.10-1.20) and larger for LDL-cholesterol (HRs > 1.40). Adding trajectory measures to reference risk models produced minimal gains in discrimination (C-index Δ +0.001-+0.003) but modest improvements in net reclassification (continuous NRI ≈ +3.0-+9.0%). Combining multiple trajectory measures from different domains (i.e. glucose, lipid and kidney function) yielded improved performance over single domain updating.

### Findings in perspective

Only a few prior large studies conducted among general populations investigated trajectory-based prediction model updating and they reported a pattern consistent with our findings. Modest added value in discrimination with small C-index gains, but measurable reclassification benefits with moderate NRIs, particularly when variability or change over time were investigated. In an individual participant meta-analysis from the Emerging Risk Factors Collaboration (191,445 persons aged 40-79), incorporating trajectory measures (cumulative mean and intercept & slope from fitted growth models) for systolic blood pressure, total cholesterol and HDL cholesterol yielded C-index increases up to +0.004 and NRI improvements of +2.4% compared with single baseline measurement models.^12^

Similarly, the Atherosclerosis Risk in Communities (ARIC) study found modest discrimination gain, with C-index increases up to +0.006 when repeated blood pressure measures were modelled as cumulative mean and intercept & slope.^13^ Larger C-index gains were reported in the Korean National Health Screening Cohort with observed C-index changes from +0.003 to +0.062 and NRIs exceeding +30% when multiple trajectory measures (average, minimum & maximum) were used simultaneously across multiple cardiometabolic measures (body mass index, systolic blood pressure, diastolic blood pressure, total cholesterol, fasting plasma glucose, smoking, and exercise).^14^

Taken together, these studies and our work suggest a consistent theme: prediction models including trajectory measures predictors, particularly those of change and dispersion such as slope, SD, IQR, min/max, tend to outperform models with only single baseline measures for cardiovascular event prediction. However, the magnitude of improvement in discrimination and risk classification is typically small and heterogeneous across populations and predictors, while gains in reclassification can be more pronounced in some settings.

Unlike the prior studies in general populations, we focused specifically on people with T2D and on three risk factors (glycemic control, atherogenic lipids, and kidney function) that are clinically relevant and routinely measured in diabetes care. We show that dynamic features of these diabetes relevant risk factors are prognostically informative, but that the absolute improvement in discrimination is limited for most adults with T2D. The relatively larger gains in older people and males mirror previous observations that baseline models tend to underperform in younger and female subgroups, leaving more room for improvement by additional predictors.

Novel circulating biomarkers such as natriuretic peptides, apolipoproteins, remnant cholesterol, or composite indices such as the triglyceride-glucose index, and polygenic risk scores have been proposed to improve CVD prediction in T2D. Published evaluations of many novel biomarkers often report small C-index increases and limited routine availability in electronic health records (EHRs), which constrains clinical implementation.^21^ Polygenic risk scores can stratify lifetime risk but so far provide incremental discrimination that is modest for short-term CVD prediction and raise implementation challenges.^22^ In contrast, trajectory measures have the advantage of being derived from routinely collected EHR data, making them pragmatic for real-world risk modeling even if their discrimination gains are modest.

### Clinical implications and future research

Our analysis demonstrates that a basic model built from routinely collected registry variables can be meaningfully augmented by trajectory metrics already present in longitudinal records, but also show that it is not yet a turnkey clinical tool. To improve performance, feasible strategies could include leveraging longer historical windows to capture earlier trajectories, incorporating a wide array of time-varying co-predictors (medication adherence, dose changes, comorbidity trajectories), and testing alternative TM definitions (e.g. time-weighted averages, variability independent of mean). Moreover, it is also important to explore effect modification: our subgroup signals (by age, sex, and medication use) indicate that the predictive value of trajectory measures may differ across sex, age groups, and treatment strata (antihypertensives, statins, glucose lowering medication). Framing future work as iterative registry-based case studies, first to identify which trajectory measures and subgroups yield the largest, reproducible gains, then to refine and externally validate models, offers a pragmatic path from exploratory findings to clinically useful prediction tools.

Methodological advances and complementary data sources offer additional avenues to increase clinical impact. Beyond simple summary trajectory measures (mean, median, SD, IQR), methods like functional data analysis, joint longitudinal-survival models, or machine learning approaches that capture nonlinear patterns, intra-individual variability, and irregular measurement timing hold promise for more detailed signal extraction from repeated measures. Recent methodological work also emphasizes quantifying longitudinal variability in change over time in prediction models which might merit evaluation in people with T2D.^23^ Finally, combining trajectory measures with targeted biomarkers or electrophysiological features (e.g. NT-proBNP or ECG metrics^24^) could produce larger, clinically meaningful improvements than any single approach alone, although for now these are not yet routinely measured.

### Strengths and limitations

This study leverages a large, nationwide registry with comprehensive longitudinal clinical registration of laboratory measurements and cardiovascular events, enabling robust estimation of trajectory measures and their associations with MACE. However, the findings derive from a Danish registry population with specific healthcare patterns, and effect sizes and implementation feasibility may differ in other health systems influencing generalizability. We evaluated multiple performance domains such as association, discrimination (C-index), and risk classification (NRI), and explicitly explored differential effects across clinically relevant subgroups (age, sex, medication use).

The inclusion of people with ≥2 measurements in the lead-up period might introduce a potential selection bias. People living with T2D with more frequent testing may differ systematically (health status, adherence to control appointment invitations, clinician concern) from those with sparse data. However, 42.6% of the excluded individuals were residents in the Southern Denmark region at baseline. Their measurements were missing due to geography, and not due to other - possibly clinically related – factors. Also, the number and timing of measurements influence trajectory measure estimates and may bias the associations. Nonetheless, this limitation is intrinsic to registry-based prediction modelling and difficult to avoid. The exclusion of relatively more healthy people living with T2D, and resulting more homogeneous case-mix could also explain the modest discriminative performance of the reference models and improvements with Trajectory measures.

Interpreted as a case study on registry-based prediction modelling, our work demonstrates practical, immediately actionable approaches for EHR-based risk modelling and identifies promising directions for refinement.

## Conclusion

Trajectory measures of dispersion or change in HbA1c, LDL-cholesterol and eGFR derived from routinely registered clinical data are associated with incident CVD in people with type 2 diabetes and moderately enhance risk classification, with small improvements in discrimination. Given their availability in electronic health records, trajectory measures might represent a pragmatic, low-cost enhancement to existing models, but their limited incremental discrimination indicates that combining trajectory measures with advanced modeling techniques and complementary risk predictors will likely be necessary to achieve clinically meaningful improvements in CVD risk prediction and management for people living with T2D.

## Colophon

## Acknowledgments

The authors gratefully acknowledge the contribution of Statistics Denmark and the Danish Health Data Authority by maintaining the Danish register infrastructure and making the data available for research. We also extend our sincere thanks to all the Danes who’s register data were used for this particular study, and to the research staff of the Steno Diabetes Center Aarhus for their invaluable contribution.

## Author contributions

PPH, OSR, JS, MTB, AAI, and DRW contributed to conception, design, data analyses, interpretation of the results, review, and editing. PPH drafted the first manuscript and all authors reviewed the subsequent versions. All authors agreed to be accountable for aspects pertaining integrity or accuracy, and approved the final version.

## Data availability

The Danish register data that support the findings of this study are owned by and available through Statistics Denmark and the Danish Health Data Authority. However, fees apply and researchers must be affiliated with an approved research institute in Denmark. Information on requesting data can be found at https://www.dst.dk/en/TilSalg/Forskningsservice.

## Ethics declaration

This study was approved by the DARTER-project steering committee at Steno Diabetes Center Aarhus, Statistics Denmark and the Danish Health Data Authority.

## List of abbreviations

ATC: Anatomical Therapeutic Chemical
ARIC: Atherosclerosis Risk in Communities
CI: Confidence Interval
CVD: Cardiovascular disease
eGFR: Estimated glomerular filtration rate
EHRs: Electronic health records
HbA1c: Glycated hemoglobin
HR: Hazard ratios
IQR: Interquartile range
LDL-C: Low-density lipoprotein cholesterol
MACE: Major Adverse Cardiovascular Events
NPU: Nomenclature for Properties and Units
NRI: Net reclassification index
RLRR: Register of Laboratory Results for Research
SD: Standard deviation
T2D: Type 2 diabetes

