## supplementary file for "Utility of glucose, lipid and kidney function Trajectory Measures for incident Cardiovascular Disease risk prediction for people living with Type 2 Diabetes: a case-study using Danish registry data"

Table S1. Definitions of variables with associated codes used in the study.

| Variables | Codes | Registers | Notes |
| --- | --- | --- | --- |
| T2D | ICD-10 codes of T2D: E10-E14, O24 (except O24.4), G623, H360, N08.3, G590, H289, H334B, T383, M142<br><br>NPU codes: NPU27300, NPU03835<br><br>ATC-codes: A10 | DNPR, RLRR, NPR | The Open-Source Diabetes Classifier defines diabetes by either: HbA1c measurements ≥48 mmol/mol (6.5%), hospital diagnosis of diabetes or prescription of glucose lowering medication. Earliest date is taken as incidence date.<br>T1D and T2D are distinguished based on hospital diagnoses of diabetes and patterns of insulin purchases. |
| HbA1c | NPU codes: NPU27300, NPU03835 | RLRR | % was converted to mmol/mol with formula: HbA1c(mmol/mol) = 10.93*HbA1c(%)-23.5 |
| LDL-cholesterol | NPU codes: NPU01568, NPU10171 | RLRR | |
| HDL-cholesterol | NPU codes: NPU01567, NPU10157 | RLRR | |
| eGFR | NPU codes: DNK35301, DNK35302, DNK35303, DNK35304, DNK35131, NPU28811, NPU28812 | RLRR | |
| Blood pressure lowering medication | ATC codes: C02, C03, C07, C08, C09 | NPR | |
| Lipid lowering medication | ATC codes: C10 | NPR | |
| Oral glucose lowering medication | ATC codes: A10B | NPR | |
| Insulin glucose lowering medication | ATC codes: A10A | NPR | |
| IHD | ICD-8 codes: 410, 411, 412, 413, 414<br><br>ICD-10 codes: I20, I21,I22, I23, I24, | DNPR, DDR | Used to determine prevalent CVD at baseline and incident CVD-events or CVD-mortality after baseline |

| | | | |
|---|---|---|---|
| | I25<br><br>SKS procedure codes:<br>KFNA-KFNG excluding KFNG20, KFNG22 | | |
| Stroke | ICD-8 codes:<br>431, 432, 433, 434, 436<br><br>ICD-10 codes:<br>I61, I63, I64, I65, I66, I693, I694<br><br>SKS procedure codes:<br>KAAL10, KAAL11, KPAQ10, KPAQ20, KPAQ21 | DNPR, DDR | Used to determine prevalent CVD at baseline and incident CVD-events or CVD-mortality after baseline |
| Heart failure | ICD-8 codes:<br>428<br><br>ICD-10 codes:<br>I110, I130, I132, I50 | DNPR, DDR | Used to determine prevalent CVD at baseline and incident CVD-events or CVD-mortality after baseline |

ATC, Anatomical Therapeutic Chemical classification; DNDR, Danish National Death registers; DNPR, The Danish National Patient Registry; eGFR, estimated glomerular filtration rate; Hb1Ac, glycosylated hemoglobin; HDL, high-density lipoprotein; ICD, International Classification of Diseases; IHD, ischemic heart disease; LDL, low-density lipoprotein; NPR, The Danish National Prescription Registry; NPU, Nomenclature for Properties and Units; RLRR, Register of Laboratory Results for Research; T1D, type 1 diabetes; T2D, type 2 diabetes.

Table S2. Baseline characteristics of study sample stratified by age group (<40/40-60/>60), sex and blood pressure lowering, lipid lowering, oral glucose lowering medication, and insulin use.

| **Characteristic** | **Age** | | | **Sex** | | **Blood pressure lowering medication** | | **Lipid lowering medication** | | **Oral glucose lowering medication** | | **Insulin** | |
|---|---|---|---|---|---|---|---|---|---|---|---|---|---|
| | **<40** | **40-60** | **>60** | **Female** | **Male** | **No** | **Yes** | **No** | **Yes** | **No** | **Yes** | **No** | **Yes** |
| Number | 2,392 | 24,845 | 56,089 | 40,122 | 43,204 | 20,055 | 63,271 | 25,575 | 57,751 | 16,493 | 66,833 | 68,597 | 14,729 |
| Age (years) | 34 (±5) | 52 (±5) | 72(±8) | 66 (±13) | 64 (±12) | 58 (±13) | 67 (±11) | 63 (±14) | 66 (±11) | 66 (±14) | 65 (±12) | 65 (±12) | 64 (±12) |
| Age at T2D diagnosis (years) | 29 (±7) | 46 (±7) | 63 (±9) | 58 (±13) | 57 (±12) | 52 (±13) | 59 (±12) | 56 (±14) | 58 (±12) | 59 (±15) | 57 (±12) | 59 (±12) | 51 (±12) |
| T2D duration (years) | 3 (2-6) | 5 (3-8) | 6 (3-11) | 6 (3-11) | 6 (3-10) | 4 (2-8) | 6 (3-11) | 5 (2-9) | 6 (3-11) | 4 (2-10) | 6 (3-11) | 5 (3-8) | 13 (8-17) |
| Female (yes) | 50% | 45% | 49% | 100% | 0% | 46% | 49% | 48% | 48% | 54% | 47% | 49% | 46% |
| HbA1C (mmol/mol) | 50 (42-62) | 50 (44-60) | 48 (43-54) | 48 (43-55) | 49 (43-57) | 48 (43-57) | 48 (43-56) | 47 (42-55) | 49 (44-56) | 45 (40-51) | 49 (44-57) | 47 (42-53) | 60 (52-70) |
| Total cholesterol (mmol/L) | 4.4 (3.8-5.2) | 4.3 (3.7-5.0) | 4.2 (3.6-4.8) | 4.4 (3.8-5.1) | 4.1 (3.5-4.7) | 4.4 (3.8-5.2) | 4.1 (3.6-4.8) | 4.8 (4.2-5.5) | 4.0 (3.5-4.6) | 4.6 (3.9-5.4) | 4.1 (3.6-4.8) | 4.3 (3.7-4.9) | 4.0 (3.5-4.7) |
| HDL-cholesterol (mmol/L) | 1.1 (0.9-1.3) | 1.2 (1.0-1.4) | 1.3 (1.1-1.6) | 1.4 (1.1-1.7) | 1.2 (1.0-1.4) | 1.2 (1.0-1.5) | 1.3 (1.0-1.5) | 1.2 (1.0-1.5) | 1.3 (1.0-1.5) | 1.3 (1.1-1.7) | 1.2 (1.0-1.5) | 1.3 (1.0-1.5) | 1.2 (1.0-1.5) |
| LDL-cholesterol (mmol/L) | 2.5 (1.9-3.1) | 2.3 (1.7-2.9) | 2.1 (1.6-2.7) | 2.2 (1.7-2.9) | 2.1 (1.6-2.7) | 2.4 (1.8-3.1) | 2.1 (1.6-2.7) | 2.7 (2.2-3.3) | 1.9 (1.5-2.4) | 2.5 (1.9-3.2) | 2.1 (1.6-2.7) | 2.2 (1.7-2.8) | 2.0 (1.5-2.5) |
| eGFR (mL/min/1.73m2) | 90 (90-90) | 90 (81-90) | 77 (62-90) | 80 (64-90) | 85 (71-90) | 90 (79-90) | 80 (64-90) | 86 (70-90) | 82 (67-90) | 80 (64-90) | 84 (69-90) | 83 (69-90) | 81 (62-90) |
| Blood pressure lowering medication use (yes) | 31% | 62% | 84% | 77% | 75% | 0% | 100% | 63% | 82% | 69% | 78% | 75% | 81% |
| Lipid lowering medication use (yes) | 40% | 64% | 73% | 69% | 69% | 52% | 75% | 0% | 100% | 50% | 74% | 68% | 75% |
| Oral glucose lowering medication use (yes) | 74% | 82% | 80% | 78% | 83% | 74% | 82% | 68% | 86% | 0% | 100% | 81% | 76% |
| Insulin medication use (yes) | 23% | 18% | 17% | 17% | 18% | 14% | 19% | 14% | 19% | 22% | 17% | 0% | 100% |

Data are presented as mean (±SD), median (Q1-Q3), or proportion (%). Baseline measurements were defined as the most recent measurement before the baseline index date of 1st of January 2015.

T2D, type 2 diabetes; HbA1c, glycated hemoglobin A1c; LDL, low-density lipoprotein; HDL, high-density lipoprotein; eGFR, estimated glomerular filtration rate.

Table S3. Summary statistics for the individual trajectory measures, along with the number of repeated measurements and their time-span of study sample stratified by age group (<40/40-60/>60), sex and blood pressure lowering, lipid lowering, oral glucose lowering medication, and insulin use.

| **Trajectory measure** | **Age** | | | **Sex** | | **Blood pressure lowering medication** | | **Lipid lowering medication** | | **Oral glucose lowering medication** | | **Insulin** | |
|---|---|---|---|---|---|---|---|---|---|---|---|---|---|
| | **<40** | **40-60** | **>60** | **Female** | **Male** | **No** | **Yes** | **No** | **Yes** | **No** | **Yes** | **No** | **Yes** |
| Number | 2,392 | 24,845 | 56,089 | 40,122 | 43,204 | 20,055 | 63,271 | 25,575 | 57,751 | 16,493 | 66,833 | 68,597 | 14,729 |
| HbA1c (mmol/mol) | | | | | | | | | | | | | |
| Number of measurements | 5 (3-8) | 5 (3-8) | 6 (4-9) | 6 (4-9) | 6 (4-9) | 5 (3-8) | 6 (4-9) | 5 (3-7) | 6 (4-9) | 5 (3-8) | 6 (4-9) | 5 (3-8) | 7 (4-10) |
| Time-span (days) | 699 (325-925) | 765 (396-951) | 849 (522-973) | 831 (489-967) | 812 (456-965) | 744 (367-942) | 844 (517-972) | 734 (360-934) | 859 (540-976) | 770 (452-944) | 834 (480-971) | 799 (447-959) | 903 (600-991) |
| mean | 53 (45-65) | 51 (45-62) | 48 (44-56) | 48 (44-56) | 50 (45-59) | 50 (44-59) | 49 (44-57) | 48 (43-57) | 50 (45-58) | 45 (41-51) | 50 (45-59) | 48 (44-54) | 62 (55-72) |
| SD | 6 (3-11) | 4 (2-9) | 3 (2-6) | 3 (2-6) | 4 (2-7) | 4 (2-8) | 3 (2-6) | 3 (2-7) | 3 (2-6) | 2 (1-4) | 4 (2-7) | 3 (2-6) | 6 (4-11) |
| median | 51 (44-63) | 50 (45-61) | 48 (44-55) | 48 (44-55) | 50 (44-58) | 49 (44-58) | 49 (44-56) | 48 (43-56) | 49 (44-57) | 45 (41-51) | 50 (45-57) | 47 (43-53) | 61 (54-71) |
| IQR | 6 (3-12) | 4 (2-9) | 3 (2-6) | 3 (2-6) | 4 (2-8) | 4 (2-8) | 4 (2-7) | 3 (2-7) | 4 (2-7) | 3 (1-5) | 4 (2-8) | 3 (2-6) | 7 (4-12) |
| intercept | 51 (43-63) | 50 (44-60) | 48 (43-54) | 48 (43-55) | 49 (44-57) | 49 (44-57) | 48 (44-56) | 48 (43-55) | 49 (44-56) | 45 (41-51) | 49 (44-57) | 47 (43-53) | 60 (52-70) |
| slope | 1 (±5) | 1 (±4) | 1 (±3) | 1 (±3) | 1 (±4) | 1 (±4) | 1 (±3) | 1 (±4) | 1 (±4) | 1 (±3) | 1 (±4) | 1 (±3) | 1 (±5) |
| LDL-cholesterol (mmol/L) | | | | | | | | | | | | | |
| Number of measurements | 3 (2-5) | 3 (2-5) | 3 (3-5) | 3 (3-5) | 3 (2-5) | 3 (2-5) | 3 (3-5) | 3 (2-5) | 4 (3-5) | 3 (2-5) | 3 (3-5) | 3 (3-5) | 3 (2-5) |
| Time-span (days) | 567 (258-805) | 659 (316-846) | 721 (379-874) | 712 (366-866) | 700 (356-861) | 657 (307-846) | 715 (373-868) | 636 (294-827) | 722 (387-877) | 693 (366-852) | 707 (359-866) | 707 (364-861) | 699 (345-868) |
| mean | 3 (±1) | 2 (±1) | 2 (±1) | 2 (±1) | 2 (±1) | 3 (±1) | 2 (±1) | 3 (±1) | 2 (±1) | 3 (±1) | 2 (±1) | 2 (±1) | 2 (±1) |
| SD | 0.4 (0.2-0.6) | 0.4 (0.2-0.6) | 0.3 (0.2-0.5) | 0.3 (0.2-0.6) | 0.3 (0.2-0.6) | 0.4 (0.2-0.6) | 0.3 (0.2-0.5) | 0.3 (0.2-0.5) | 0.3 (0.2-0.6) | 0.3 (0.2-0.6) | 0.3 (0.2-0.6) | 0.3 (0.2-0.6) | 0.3 (0.2-0.5) |
| median | 3 (±1) | 2 (±1) | 2 (±1) | 2 (±1) | 2 (±1) | 3 (±1) | 2 (±1) | 3 (±1) | 2 (±1) | 3 (±1) | 2 (±1) | 2 (±1) | 2 (±1) |
| IQR | 0.4 (0.2-0.6) | 0.3 (0.2-0.6) | 0.3 (0.2-0.5) | 0.3 (0.2-0.5) | 0.3 (0.2-0.5) | 0.3 (0.2-0.6) | 0.3 (0.2-0.5) | 0.3 (0.2-0.5) | 0.3 (0.2-0.6) | 0.3 (0.2-0.5) | 0.3 (0.2-0.5) | 0.3 (0.2-0.5) | 0.3 (0.2-0.5) |
| intercept | 2 (±1) | 2 (±1) | 2 (±1) | 2 (±1) | 2 (±1) | 2 (±1) | 2 (±1) | 3 (±1) | 2 (±1) | 2 (±1) | 2 (±1) | 2 (±1) | 2 (±1) |
| slope | 0.1 (±0.1) | 0.1 (±0.1) | 0.1 (±0.1) | 0.1 (±0.1) | 0.1 (±0.1) | 0.1 (±0.1) | 0.1 (±0.1) | 0.1 (±0.1) | 0.1 (±0.1) | 0.1 (±0.1) | 0.1 (±0.1) | 0.1 (±0.1) | 0.1 (±0.1) |
| eGFR (mL/min/1.73m$^2$) | | | | | | | | | | | | | |

| | | | | | | | | | | | | | |
|---|---|---|---|---|---|---|---|---|---|---|---|---|---|
| Number of measurements | 5 (3-8) | 5 (3-8) | 6 (4-10) | 6 (4-10) | 5 (3-9) | 5 (3-8) | 6 (4-10) | 5 (3-9) | 6 (4-9) | 6 (4-10) | 6 (3-9) | 5 (3-9) | 6 (4-11) |
| Time-span (days) | 702 (348-906) | 754 (438-934) | 823 (572-965) | 813 (551-959) | 787 (496-952) | 742 (406-927) | 819 (562-964) | 763 (432-942) | 815 (560-961) | 809 (567-960) | 796 (509-955) | 791 (512-952) | 836 (561-973) |
| mean | 90 (90-99) | 90 (83-93) | 78 (65-88) | 81 (67-90) | 85 (73-90) | 89 (80-92) | 81 (67-90) | 85 (72-90) | 83 (69-90) | 81 (65-90) | 84 (71-90) | 84 (71-90) | 82 (65-90) |
| SD | 6 (0-12) | 6 (2-10) | 6 (3-8) | 6 (3-9) | 5 (3-9) | 5 (2-9) | 6 (3-9) | 6 (3-9) | 6 (3-9) | 6 (3-9) | 6 (3-9) | 6 (3-9) | 6 (3-9) |
| median | 90 (90-90) | 90 (83-90) | 78 (65-90) | 81 (66-90) | 86 (73-90) | 90 (80-90) | 81 (67-90) | 86 (72-90) | 83 (69-90) | 81 (65-90) | 85 (71-90) | 84 (71-90) | 83 (64-90) |
| IQR | 3 (0-11) | 5 (0-10) | 6 (3-9) | 6 (3-10) | 5 (2-9) | 5 (1-9) | 6 (3-9) | 5 (2-9) | 6 (2-9) | 6 (3-9) | 5 (2-9) | 5 (2-9) | 5 (2-10) |
| intercept | 89 (87-91) | 88 (81-89) | 77 (63-87) | 80 (65-88) | 84 (71-89) | 87 (79-89) | 79 (65-88) | 84 (71-89) | 81 (67-88) | 79 (64-88) | 83 (69-89) | 82 (69-89) | 80 (63-88) |
| slope | 2 (±4) | 1 (±3) | 1 (±3) | 1 (±3) | 1 (±3) | 1 (±3) | 1 (±3) | 1 (±3) | 1 (±3) | 1 (±3) | 1 (±3) | 1 (±3) | 2 (±4) |

Data are presented as mean (±SD), median (Q1-Q3), frequencies (n) or time (days). The trajectory measures were calculated including all recorded measurements within the 3 years prior to the baseline index date of 1st of January 2015. (2012, 2013 and 2014).

Note: the trajectory measures were paired as mean & SD, median & IQR and intercept & slope from a fitted growth model.

T2D: type 2 diabetes; HbA1c: glycated hemoglobin A1c; LDL: low-density lipoprotein; eGFR: estimated glomerular filtration rate; SD: Standard deviation, IQR: Interquartile range.

Table S4. Incidence rates per 1000 person-years of MACE endpoint for total study sample, and stratified by sex, age group and blood pressure lowering, lipid lowering, oral glucose lowering medication, and insulin use.

| **Characteristic** | **Stratum** | **Events** | **Person-years** | **Incidence rate (95%CI) per 1000 person-years** |
|---|---|---|---|---|
| Total | | 11280 | 443130 | 25.5 (25-25.9) |
| Age (years) | <40 | 62 | 14096 | 4.4 (3.4-5.6) |
| | 40-60 | 2108 | 140729 | 15 (14.3-15.6) |
| | >60 | 9110 | 288305 | 31.6 (31-32.3) |
| Sex | Male | 6637 | 226992 | 29.2 (28.5-30) |
| | Female | 4643 | 216138 | 21.5 (20.9-22.1) |
| Blood pressure lowering medication | No | 1871 | 111240 | 16.8 (16.1-17.6) |
| | Yes | 9409 | 331890 | 28.3 (27.8-28.9) |
| Lipid lowering medication | No | 3658 | 133878 | 27.3 (26.4-28.2) |
| | Yes | 7622 | 309252 | 24.6 (24.1-25.2) |
| Oral glucose lowering medication | No | 2412 | 85331 | 28.3 (27.1-29.4) |
| | Yes | 8868 | 357799 | 24.8 (24.3-25.3) |
| Insulin | No | 8698 | 367861 | 23.6 (23.2-24.1) |
| | Yes | 2582 | 75268 | 34.3 (33-35.7) |

Data are presented as Incidence rate (95%CI) per 1000 person-years for MACE: Ischemic heart disease, stroke, heart failure or CVD-mortality. CI, Confidence Interval.

Table S5. Association of paired trajectory measures with MACE endpoint, after adjustment for baseline predictors, stratified by age group (<40/40-60/>60), sex and blood pressure lowering, lipid lowering, oral glucose lowering medication, and insulin use.

| Trajectory measure | Adjusted HR (95%CI) for MACE | | | | | | | | | | | | |
|---|---|---|---|---|---|---|---|---|---|---|---|---|---|
| | Age | | | Sex | | Blood pressure lowering medication | | Lipid lowering medication | | Oral glucose lowering medication | | Insulin | |
| | **<40** | **40-60** | **>60** | **Female** | **Male** | **No** | **Yes** | **No** | **Yes** | **No** | **Yes** | **No** | **Yes** |
| HbA1c[1] | | | | | | | | | | | | | |
| mean | 1.04 (0.90-1.21) | 1.0.3 (1.00-1.06) | 0.99 (0.97-1.02) | 1.00 (0.97-1.03) | 1.01 (0.99-1.04) | 1.02 (0.98-1.05) | 1.00 (0.98-1.02) | 1.02 (0.99-1.05) | 1.00 (0.98-1.02) | 0.98 (0.94-1.01) | 1.02 (0.99-1.04) | 1.00 (0.97-1.02) | 1.04 (1.00-1.07) |
| SD | 0.93 (0.63-1.36) | 1.01 (0.93-1.08) | 1.12 (1.07-1.18) | 1.11 (1.04-1.19) | 1.06 (1.01-1.11) | 1.04 (0.97-1.13) | 1.09 (1.04-1.14) | 1.05 (0.98-1.11) | 1.10 (1.04-1.16) | 1.19 (1.08-1.30) | 1.06 (1.02-1.11) | 1.10 (1.05-1.16) | 1.06 (0.99-1.13) |
| median | 1.05 (0.92-1.20) | 1.02 (1.00-1.05) | 1.01 (0.99-1.03) | 1.01 (0.98-1.04) | 1.02 (0.99-1.04) | 1.02 (0.99-1.06) | 1.01 (0.99-1.03) | 1.02 (1.00-1.05) | 1.01 (0.99-1.03) | 0.99 (0.96-1.02) | 1.02 (1.00-1.04) | 1.01 (0.98-1.03) | 1.04 (1.01-1.07) |
| IQR | 0.92 (0.69-1.23) | 1.03 (0.98-1.09) | 1.07 (1.03-1.11) | 1.06 (1.01-1.12) | 1.05 (1.01-1.09) | 1.03 (0.97-1.09) | 1.06 (1.03-1.10) | 1.03 (0.98-1.08) | 1.07 (1.03-1.11) | 1.10 (1.02-1.19) | 1.05 (1.01-1.08) | 1.07 (1.03-1.11) | 1.05 (1.00-1.10) |
| intercept | 1.04 (0.90-1.20) | 1.03 (1.00-1.06) | 1.02 (1.00-1.04) | 1.02 (1.00-1.05) | 1.03 (1.01-1.05) | 1.02 (0.99-1.06) | 1.02 (1.01-1.04) | 1.03 (1.00-1.05) | 1.02 (1.00-1.05) | 1.01 (0.98-1.04) | 1.03 (1.01-1.05) | 1.03 (1.00-1.05) | 1.05 (1.02-1.08) |
| slope | 1.03 (0.62-1.70) | 1.14 (1.03-1.26) | 1.17 (1.10-1.25) | 1.16 (1.06-1.27) | 1.17 (1.09-1.25) | 1.24 (1.11-1.38) | 1.14 (1.07-1.21) | 1.15 (1.05-1.26) | 1.17 (1.09-1.25) | 1.14 (0.99-1.31) | 1.17 (1.11-1.25) | 1.18 (1.10-1.27) | 1.18 (1.08-1.29) |
| LDL-cholesterol[1] | | | | | | | | | | | | | |
| mean | 1.07 (0.8-1.43) | 1.02 (0.97-1.08) | 1.01 (0.98-1.03) | 1.01 (0.98-1.05) | 1.01 (0.98-1.04) | 1.04 (0.99-1.10) | 1.01 (0.98-1.03) | 1.01 (0.97-1.06) | 1.01 (0.97-1.04) | 1.00 (0.96-1.05) | 1.01 (0.98-1.04) | 1.01 (0.98-1.03) | 1.03 (0.97-1.08) |
| SD | 1.49 (1.05-2.12) | 1.28 (1.14-1.44) | 1.10 (1.03-1.17) | 1.14 (1.05-1.24) | 1.15 (1.07-1.234) | 1.09 (0.96-1.24) | 1.16 (1.09-1.23) | 1.17 (1.04-1.30) | 1.14 (1.07-1.22) | 1.14 (1.01-1.28) | 1.15 (1.08-1.22) | 1.16 (1.09-1.23) | 1.08 (0.95-1.22) |
| median | 1.10 (0.84-1.43) | 1.02 (0.97-1.08) | 1.01 (0.99-1.04) | 1.01 (0.98-1.05) | 1.01 (0.98-1.05) | 1.04 (0.99-1.09) | 1.01 (0.98-1.03) | 1.02 (0.98-1.06) | 1.01 (0.98-1.05) | 1.01 (0.96-1.06) | 1.02 (0.99-1.04) | 1.01 (0.99-1.04) | 1.02 (0.97-1.08) |
| IQR | 1.68 (1.14-2.49) | 1.20 (1.08-1.33) | 1.09 (1.03-1.15) | 1.10 (1.03-1.19) | 1.12 (1.05-1.20) | 1.07 (0.96-1.19) | 1.128 (1.069-1.190) | 1.16 (1.05-1.29) | 1.10 (1.04-1.17) | 1.08 (0.98-1.20) | 1.12 (1.06-1.19) | 1.12 (1.06-1.18) | 1.07 (0.96-1.20) |
| intercept | 1.24 (0.90- | 1.07 (1.01- | 1.03 (1.00- | 1.04 (1.00- | 1.04 (1.00- | 1.05 (0.99- | 1.04 (1.01-1.07) | 1.03 (0.98- | 1.05 (1.01- | 1.02 (0.97- | 1.04 (1.01- | 1.03 (1.00- | 1.06 (1.00- |

| | | | | | | | | | | | | | |
|---|---|---|---|---|---|---|---|---|---|---|---|---|---|
| | 1.72) | 1.13) | 1.06) | 1.08) | 1.08) | 1.12) | | 1.08) | 1.08) | 1.08) | 1.08) | 1.07) | 1.12) |
| slope | 2.39 (0.37-15.4) | 1.83 (1.27-2.64) | 1.37 (1.13-1.67) | 1.54 (1.19-1.99) | 1.42 (1.13-1.79) | 2.02 (1.35-3.02) | 1.38 (1.14-1.67) | 1.16 (0.82-1.64) | 1.58 (1.30-1.92) | 1.35 (0.94-1.95) | 1.52 (1.25-1.84) | 1.44 (1.19-1.75) | 1.69 (1.17-2.44) |
| eGFR[1] | | | | | | | | | | | | | |
| mean | 0.95 (0.80-1.13) | 0.98 (0.95-1.01) | 1.00 (0.98-1.01) | 0.99 (0.98-1.01) | 0.99 (0.98-1.01) | 0.99 (0.96-1.03) | 0.99 (0.98-1.01) | 1.00 (0.98-1.01) | 0.99 (0.98-1.01) | 0.99 (0.98-1.01) | 0.99 (0.98-1.01) | 1.00 (0.98-1.01) | 0.99 (0.97-1.01) |
| SD | 1.43 (1.01-2.01) | 1.17 (1.09-1.26) | 1.21 (1.16-1.26) | 1.21 (1.14-1.28) | 1.20 (1.14-1.26) | 1.14 (1.05-1.25) | 1.21 (1.16-1.26) | 1.15 (1.08-1.23) | 1.22 (1.17-1.28) | 1.25 (1.15-1.36) | 1.18 (1.13-1.24) | 1.22 (1.17-1.28) | 1.12 (1.04-1.21) |
| median | 0.97 (0.82-1.14) | 0.98 (0.95-1.01) | 1.00 (0.99-1.01) | 1.00 (0.98-1.01) | 1.00 (0.99-1.01) | 1.00 (0.97-1.03) | 1.00 (0.99-1.01) | 1.00 (0.98-1.02) | 1.00 (0.99-1.01) | 1.00 (0.98-1.02) | 1.00 (0.99-1.01) | 1.00 (0.99-1.01) | 0.99 (0.98-1.01) |
| IQR | 1.13 (0.89-1.44) | 1.10 (1.04-1.15) | 1.13 (1.10-1.17) | 1.12 (1.08-1.17) | 1.12 (1.09-1.16) | 1.09 (1.02-1.16) | 1.13 (1.09-1.16) | 1.08 (1.03-1.13) | 1.15 (1.11-1.18) | 1.17 (1.11-1.23) | 1.11 (1.07-1.14) | 1.13 (1.10-1.17) | 1.08 (1.03-1.14) |
| intercept | 1.02 (0.84-1.23) | 0.99 (0.96-1.02) | 1.00 (0.99-1.02) | 1.00 (0.99-1.02) | 1.00 (0.99-1.02) | 1.01 (0.97-1.04) | 1.00 (0.99-1.01) | 1.00 (0.98-1.02) | 1.00 (0.99-1.02) | 1.00 (0.98-1.02) | 1.00 (0.99-1.02) | 1.01 (0.99-1.02) | 1.00 (0.98-1.02) |
| slope | 1.30 (0.66-2.54) | 1.09 (0.95-1.25) | 1.09 (1.01-1.17) | 1.12 (1.01-1.23) | 1.08 (0.99-1.18) | 1.19 (1.02-1.40) | 1.08 (1.01-1.16) | 1.05 (0.94-1.17) | 1.12 (1.03-1.21) | 1.17 (1.03-1.34) | 1.07 (1.00-1.16) | 1.11 (1.03-1.20) | 1.08 (0.96-1.21) |

[1] HbA1c: per 10 mmol/mol, LDL-cholesterol: per 1 mmol/l, eGFR: per 10 ml/min/1,73m2.
Data are presented as HR (95%CI) for MACE: Ischemic heart disease, stroke, heart failure or CVD-mortality.
Note: The trajectory measures were calculated including all recorded measurements within the 3 years prior to the baseline index date of 1st of January 2015. (2012, 2013 and 2014) and paired as mean & SD, median & IQR, and intercept & slope from a fitted growth model. The trajectory measure models were fitted using an offset procedure, sequentially adding one of the three types of paired trajectory measures to the respective (sub)sample reference model: mean & SD, median & IQR, and intercept & slope.  All reference models (Cox-regression) included baseline measurements for age, age at T2D diagnosis, sex, HbA1c, LDL-cholesterol, HDL-cholesterol, eGFR, blood pressure lowering, lipid lowering, oral glucose lowering medication and insulin use.
HbA1c: Hemoglobulin A1c, LDL: low density lipoprotein, eGFR: estimated glomerular filtration rate, HR: Hazard ratio, CI: Confidence Interval, SD: Standard deviation, IQR: Interquartile range.

Table S6. The baseline reference models' coefficients and HRs for total study sample, and stratified by sex, age group and blood pressure lowering, lipid lowering, oral glucose lowering medication, and insulin use.

| **Stratum & Predictor** | **Coefficient (95%CI)** | **HR (95%CI)** |
|---|---|---|
| Total Sample | | |
| Age | 0.057 (0.054, 0.061) | 1.059 (1.055-1.063) |
| Age at T2D diagnosis | -0.017 (-0.021, -0.014) | 0.983 (0.979-0.986) |
| Female | 0.403 (0.363, 0.442) | 1.496 (1.438-1.556) |
| HbA1c | 0.076 (0.061, 0.092) | 1.079 (1.062-1.097) |
| LDL-cholesterol | 0.125 (0.102, 0.148) | 1.133 (1.107-1.159) |
| HDL-cholesterol | -0.187 (-0.237, -0.137) | 0.830 (0.789-0.872) |
| eGFR | -0.053 (-0.064, -0.043) | 0.948 (0.938-0.958) |
| Blood pressure lowering medication use | 0.214 (0.162, 0.267) | 1.239 (1.176-1.306) |
| lipid lowering medication use | -0.147 (-0.192, -0.103) | 0.863 (0.826-0.902) |
| oral glucose lowering medication use | -0.061 (-0.109, -0.014) | 0.940 (0.897-0.986) |
| insulin medication use | 0.165 (0.110, 0.221) | 1.180 (1.116-1.247) |
| Age <40 | | |
| Age | 0.118 (0.044, 0.191) | 1.125 (1.045-1.211) |
| Age at T2D diagnosis | -0.073 (-0.119, -0.027) | 0.930 (0.888-0.974) |
| Female | 0.177 (-0.347, 0.701) | 1.194 (0.707-2.016) |
| HbA1c | 0.099 (-0.038, 0.236) | 1.105 (0.963-1.267) |
| LDL-cholesterol | 0.228 (-0.016, 0.472) | 1.256 (0.984-1.604) |
| HDL-cholesterol | -0.070 (-0.859, 0.719) | 0.932 (0.423-2.053) |
| eGFR | -0.017 (-0.184, 0.151) | 0.983 (0.832-1.163) |
| Blood pressure lowering medication use | 0.453 (-0.075, 0.982) | 1.574 (0.928-2.669) |
| lipid lowering medication use | 0.230 (-0.310, 0.770) | 1.258 (0.733-2.159) |
| oral glucose lowering medication use | 0.265 (-0.416, 0.946) | 1.304 (0.660-2.576) |
| insulin medication use | -0.626 (-1.361, 0.109) | 0.535 (0.256-1.116) |
| Age 40-60 | | |
| Age | 0.056 (0.044, 0.068) | 1.057 (1.045-1.070) |
| Age at T2D diagnosis | -0.022 (-0.031, -0.014) | 0.978 (0.970-0.986) |
| Female | 0.449 (0.356, 0.542) | 1.566 (1.427-1.719) |
| HbA1c | 0.095 (0.066, 0.124) | 1.100 (1.069-1.132) |
| LDL-cholesterol | 0.208 (0.160, 0.256) | 1.231 (1.173-1.291) |
| HDL-cholesterol | -0.345 (-0.481, -0.210) | 0.708 (0.618-0.810) |
| eGFR | -0.073 (-0.100, -0.045) | 0.930 (0.905-0.956) |
| Blood pressure lowering medication use | 0.216 (0.119, 0.313) | 1.241 (1.127-1.367) |
| lipid lowering medication use | -0.076 (-0.173, 0.021) | 0.927 (0.841-1.021) |
| oral glucose lowering medication use | -0.195 (-0.308, -0.081) | 0.823 (0.735-0.922) |
| insulin medication use | 0.222 (0.102, 0.342) | 1.249 (1.107-1.408) |
| Age >60 | | |
| Age | 0.058 (0.054, 0.063) | 1.060 (1.055-1.065) |
| Age at T2D diagnosis | -0.016 (-0.020, -0.012) | 0.984 (0.980-0.988) |
| Female | 0.390 (0.346, 0.434) | 1.477 (1.413-1.543) |
| HbA1c | 0.063 (0.044, 0.082) | 1.065 (1.045-1.086) |
| LDL-cholesterol | 0.097 (0.071, 0.124) | 1.102 (1.074-1.132) |
| HDL-cholesterol | -0.170 (-0.224, -0.116) | 0.844 (0.799-0.891) |
| eGFR | -0.051 (-0.062, -0.039) | 0.950 (0.940-0.961) |

| | | |
|---|---|---|
| Blood pressure lowering medication use | 0.212 (0.149, 0.276) | 1.236 (1.160-1.317) |
| lipid lowering medication use | -0.173 (-0.224, -0.123) | 0.841 (0.799-0.885) |
| oral glucose lowering medication use | -0.030 (-0.083, 0.023) | 0.970 (0.920-1.023) |
| insulin medication use | 0.168 (0.106, 0.231) | 1.183 (1.111-1.260) |
| Female | | |
| Age | 0.058 (0.052, 0.063) | 1.059 (1.053-1.066) |
| Age at T2D diagnosis | -0.014 (-0.020, -0.009) | 0.986 (0.980-0.991) |
| Female | n.a. | 0.000 () |
| HbA1c | 0.088 (0.062, 0.113) | 1.092 (1.064-1.120) |
| LDL-cholesterol | 0.101 (0.066, 0.135) | 1.106 (1.069-1.144) |
| HDL-cholesterol | -0.227 (-0.299, -0.154) | 0.797 (0.741-0.858) |
| eGFR | -0.054 (-0.070, -0.039) | 0.947 (0.932-0.962) |
| Blood pressure lowering medication use | 0.299 (0.209, 0.389) | 1.349 (1.233-1.475) |
| lipid lowering medication use | -0.189 (-0.259, -0.118) | 0.828 (0.772-0.889) |
| oral glucose lowering medication use | 0.011 (-0.061, 0.083) | 1.011 (0.941-1.086) |
| insulin medication use | 0.145 (0.056, 0.234) | 1.156 (1.057-1.263) |
| Male | | |
| Age | 0.058 (0.052, 0.063) | 1.059 (1.054-1.065) |
| Age at T2D diagnosis | -0.020 (-0.025, -0.015) | 0.981 (0.976-0.985) |
| Female | n.a. | n.a. |
| HbA1c | 0.067 (0.047, 0.087) | 1.070 (1.048-1.091) |
| LDL-cholesterol | 0.142 (0.111, 0.173) | 1.152 (1.117-1.189) |
| HDL-cholesterol | -0.152 (-0.221, -0.083) | 0.859 (0.801-0.921) |
| eGFR | -0.051 (-0.066, -0.037) | 0.950 (0.936-0.963) |
| Blood pressure lowering medication use | 0.169 (0.104, 0.234) | 1.184 (1.109-1.264) |
| lipid lowering medication use | -0.121 (-0.178, -0.064) | 0.886 (0.837-0.938) |
| oral glucose lowering medication use | -0.117 (-0.181, -0.053) | 0.890 (0.835-0.948) |
| insulin medication use | 0.176 (0.106, 0.247) | 1.193 (1.111-1.280) |
| Blood pressure lowering medication = No | | |
| Age | 0.060 (0.051, 0.070) | 1.062 (1.052-1.072) |
| Age at T2D diagnosis | -0.015 (-0.024, -0.005) | 0.985 (0.976-0.995) |
| Female | 0.499 (0.398, 0.600) | 1.647 (1.489-1.822) |
| HbA1c | 0.118 (0.086, 0.151) | 1.126 (1.090-1.163) |
| LDL-cholesterol | 0.151 (0.099, 0.202) | 1.162 (1.104-1.224) |
| HDL-cholesterol | -0.327 (-0.456, -0.198) | 0.721 (0.634-0.820) |
| eGFR | 0.002 (-0.030, 0.033) | 1.002 (0.971-1.033) |
| Blood pressure lowering medication use | n.a. | n.a. |
| lipid lowering medication use | -0.126 (-0.225, -0.026) | 0.882 (0.798-0.974) |
| oral glucose lowering medication use | -0.077 (-0.186, 0.032) | 0.926 (0.830-1.033) |
| insulin medication use | 0.124 (-0.020, 0.268) | 1.132 (0.980-1.308) |
| Blood pressure lowering medication = Yes | | |
| Age | 0.057 (0.052, 0.061) | 1.058 (1.054-1.063) |
| Age at T2D diagnosis | -0.018 (-0.022, -0.014) | 0.982 (0.978-0.986) |
| Female | 0.383 (0.339, 0.426) | 1.466 (1.404-1.531) |
| HbA1c | 0.062 (0.044, 0.080) | 1.064 (1.045-1.084) |
| LDL-cholesterol | 0.117 (0.091, 0.143) | 1.124 (1.096-1.153) |
| HDL-cholesterol | -0.158 (-0.212, -0.103) | 0.854 (0.809-0.902) |
| eGFR | -0.061 (-0.072, -0.050) | 0.941 (0.930-0.951) |

| | | |
|---|---|---|
| Blood pressure lowering medication use | n.a. | n.a. |
| lipid lowering medication use | -0.155 (-0.205, -0.105) | 0.856 (0.815-0.900) |
| oral glucose lowering medication use | -0.057 (-0.111, -0.004) | 0.944 (0.895-0.996) |
| insulin medication use | 0.180 (0.120, 0.240) | 1.197 (1.127-1.271) |
| Lipid lowering medication = No | | |
| Age | 0.060 (0.054, 0.067) | 1.062 (1.055-1.069) |
| Age at T2D diagnosis | -0.018 (-0.025, -0.012) | 0.982 (0.976-0.988) |
| Female | 0.398 (0.328, 0.467) | 1.488 (1.388-1.596) |
| HbA1c | 0.106 (0.081, 0.132) | 1.112 (1.084-1.141) |
| LDL-cholesterol | 0.126 (0.087, 0.165) | 1.134 (1.091-1.179) |
| HDL-cholesterol | -0.205 (-0.292, -0.118) | 0.815 (0.747-0.889) |
| eGFR | -0.036 (-0.055, -0.017) | 0.965 (0.947-0.983) |
| Blood pressure lowering medication use | 0.227 (0.148, 0.305) | 1.254 (1.159-1.357) |
| lipid lowering medication use | n.a. | n.a. |
| oral glucose lowering medication use | -0.113 (-0.185, -0.040) | 0.894 (0.831-0.961) |
| insulin medication use | 0.111 (0.009, 0.214) | 1.118 (1.009-1.238) |
| Lipid lowering medication = Yes | | |
| Age | 0.056 (0.051, 0.061) | 1.058 (1.052-1.063) |
| Age at T2D diagnosis | -0.017 (-0.022, -0.012) | 0.983 (0.979-0.988) |
| Female | 0.406 (0.358, 0.454) | 1.501 (1.431-1.575) |
| HbA1c | 0.059 (0.039, 0.079) | 1.061 (1.039-1.082) |
| LDL-cholesterol | 0.124 (0.095, 0.153) | 1.132 (1.100-1.165) |
| HDL-cholesterol | -0.177 (-0.239, -0.116) | 0.837 (0.787-0.891) |
| eGFR | -0.062 (-0.075, -0.049) | 0.940 (0.928-0.952) |
| Blood pressure lowering medication use | 0.211 (0.140, 0.282) | 1.235 (1.150-1.326) |
| lipid lowering medication use | n.a. | n.a. |
| oral glucose lowering medication use | -0.027 (-0.090, 0.037) | 0.974 (0.913-1.038) |
| insulin medication use | 0.192 (0.126, 0.258) | 1.211 (1.134-1.294) |
| Oral glucose lowering medication = No | | |
| Age | 0.059 (0.050, 0.067) | 1.060 (1.052-1.069) |
| Age at T2D diagnosis | -0.023 (-0.030, -0.015) | 0.977 (0.970-0.985) |
| Female | 0.521 (0.437, 0.606) | 1.684 (1.548-1.833) |
| HbA1c | 0.139 (0.102, 0.175) | 1.149 (1.108-1.192) |
| LDL-cholesterol | 0.076 (0.030, 0.123) | 1.079 (1.030-1.131) |
| HDL-cholesterol | -0.136 (-0.232, -0.041) | 0.873 (0.793-0.960) |
| eGFR | -0.079 (-0.100, -0.059) | 0.924 (0.905-0.943) |
| Blood pressure lowering medication use | 0.284 (0.176, 0.391) | 1.328 (1.193-1.478) |
| lipid lowering medication use | -0.224 (-0.314, -0.134) | 0.799 (0.731-0.874) |
| oral glucose lowering medication use | n.a. | n.a. |
| insulin medication use | -0.149 (-0.289, -0.008) | 0.862 (0.749-0.992) |
| Oral glucose lowering medication = Yes | | |
| Age | 0.058 (0.054, 0.063) | 1.060 (1.055-1.065) |
| Age at T2D diagnosis | -0.017 (-0.021, -0.012) | 0.984 (0.979-0.988) |
| Female | 0.371 (0.326, 0.416) | 1.449 (1.386-1.515) |
| HbA1c | 0.066 (0.049, 0.084) | 1.069 (1.050-1.088) |
| LDL-cholesterol | 0.139 (0.113, 0.166) | 1.149 (1.120-1.180) |
| HDL-cholesterol | -0.203 (-0.262, -0.144) | 0.816 (0.769-0.866) |
| eGFR | -0.045 (-0.057, -0.033) | 0.956 (0.944-0.968) |

| | | |
|---|---|---|
| Blood pressure lowering medication use | 0.198 (0.138, 0.259) | 1.219 (1.148-1.295) |
| lipid lowering medication use | -0.122 (-0.173, -0.070) | 0.886 (0.841-0.932) |
| oral glucose lowering medication use | n.a. | n.a. |
| insulin medication use | 0.219 (0.158, 0.279) | 1.244 (1.171-1.322) |
| Insulin = No | | |
| Age | 0.059 (0.054, 0.064) | 1.061 (1.056-1.066) |
| Age at T2D diagnosis | -0.017 (-0.021, -0.012) | 0.984 (0.979-0.988) |
| Female | 0.402 (0.357, 0.447) | 1.495 (1.429-1.564) |
| HbA1c | 0.075 (0.055, 0.095) | 1.078 (1.056-1.100) |
| LDL-cholesterol | 0.114 (0.088, 0.140) | 1.121 (1.092-1.151) |
| HDL-cholesterol | -0.145 (-0.202, -0.087) | 0.865 (0.817-0.916) |
| eGFR | -0.038 (-0.050, -0.026) | 0.963 (0.951-0.975) |
| Blood pressure lowering medication use | 0.209 (0.151, 0.266) | 1.232 (1.163-1.305) |
| lipid lowering medication use | -0.151 (-0.201, -0.101) | 0.860 (0.818-0.904) |
| oral glucose lowering medication use | -0.089 (-0.146, -0.033) | 0.915 (0.865-0.968) |
| insulin medication use | n.a. | n.a. |
| Insulin = Yes | | |
| Age | 0.053 (0.046, 0.060) | 1.055 (1.047-1.062) |
| Age at T2D diagnosis | -0.021 (-0.028, -0.015) | 0.979 (0.972-0.985) |
| Female | 0.417 (0.334, 0.500) | 1.518 (1.397-1.649) |
| HbA1c | 0.082 (0.056, 0.108) | 1.086 (1.058-1.114) |
| LDL-cholesterol | 0.151 (0.104, 0.198) | 1.162 (1.109-1.218) |
| HDL-cholesterol | -0.325 (-0.431, -0.219) | 0.722 (0.650-0.803) |
| eGFR | -0.095 (-0.116, -0.075) | 0.909 (0.891-0.928) |
| Blood pressure lowering medication use | 0.219 (0.092, 0.346) | 1.244 (1.096-1.413) |
| lipid lowering medication use | -0.150 (-0.247, -0.054) | 0.860 (0.781-0.948) |
| oral glucose lowering medication use | 0.041 (-0.053, 0.135) | 1.042 (0.948-1.145) |
| insulin medication use | n.a. | n.a. |

[1] HbA1c: per 10 mmol/mol, LDL-cholesterol: per 1 mmol/l, eGFR: per 10 ml/min/1,73m2.

Data are presented as coefficient(i.e. log(HR)) (95%CI) or HR (95%CI) for MACE: Ischemic heart disease, stroke, heart failure or CVD-mortality.

HbA1c: Hemoglobulin A1c, LDL: low density lipoprotein, eGFR: estimated glomerular filtration rate, HR: Hazard ratio, CI: Confidence Interval.

Table S7. Performance statistics of reference and trajectory measure updated prediction models.

| Metric & Prediction model | Total sample | Age | | | Sex | | Blood pressure lowering medication | | Lipid lowering medication | | Oral glucose lowering medication | | Insulin | |
|---|---|---|---|---|---|---|---|---|---|---|---|---|---|---|
| | | <40 | 40-60 | >60 | Female | Male | No | Yes | No | Yes | No | Yes | No | Yes |
| **C-index (95%CI)** | | | | | | | | | | | | | | |
| Reference | 0.665 (0.660-0.670) | 0.648 (0.582-0.715) | 0.634 (0.622-0.646) | 0.632 (0.626-0.638) | 0.675 (0.667-0.683) | 0.648 (0.642-0.655) | 0.681 (0.669-0.693) | 0.650 (0.645-0.656) | 0.688 (0.680-0.696) | 0.653 (0.647-0.659) | 0.678 (0.668-0.689) | 0.662 (0.657-0.668) | 0.660 (0.654-0.665) | 0.663 (0.653-0.673) |
| HbA1c | | | | | | | | | | | | | | |
| Mean & SD | 0.666 (0.661-0.671) | 0.649 (0.583-0.715) | 0.635 (0.623-0.647) | 0.633 (0.627-0.639) | 0.676 (0.668-0.683) | 0.649 (0.643-0.656) | 0.681 (0.669-0.693) | 0.651 (0.646-0.656) | 0.689 (0.680-0.697) | 0.654 (0.648-0.660) | 0.680 (0.670-0.690) | 0.663 (0.657-0.668) | 0.661 (0.655-0.666) | 0.665 (0.655-0.675) |
| Median & IQR | 0.666 (0.661-0.671) | 0.648 (0.583-0.714) | 0.635 (0.623-0.647) | 0.633 (0.627-0.638) | 0.675 (0.668-0.683) | 0.649 (0.643-0.656) | 0.681 (0.669-0.693) | 0.651 (0.646-0.657) | 0.689 (0.680-0.697) | 0.654 (0.648-0.660) | 0.679 (0.669-0.690) | 0.663 (0.657-0.668) | 0.660 (0.655-0.666) | 0.665 (0.655-0.675) |
| Intercept & slope | 0.666 (0.661-0.671) | 0.649 (0.583-0.714) | 0.635 (0.623-0.647) | 0.633 (0.627-0.638) | 0.675 (0.668-0.683) | 0.649 (0.643-0.656) | 0.682 (0.670-0.694) | 0.651 (0.645-0.656) | 0.689 (0.680-0.697) | 0.654 (0.647-0.660) | 0.678 (0.668-0.689) | 0.663 (0.657-0.668) | 0.661 (0.655-0.666) | 0.664 (0.654-0.674) |
| LDL-cholesterol | | | | | | | | | | | | | | |
| Mean & SD | 0.666 (0.661-0.671) | 0.670 (0.603-0.737) | 0.637 (0.625-0.649) | 0.632 (0.627-0.638) | 0.676 (0.668-0.683) | 0.649 (0.643-0.656) | 0.681 (0.670-0.693) | 0.651 (0.646-0.657) | 0.689 (0.680-0.697) | 0.654 (0.648-0.660) | 0.679 (0.669-0.689) | 0.663 (0.657-0.668) | 0.661 (0.655-0.666) | 0.664 (0.654-0.674) |
| Median & IQR | 0.666 (0.661-0.671) | 0.686 (0.617-0.755) | 0.636 (0.624-0.648) | 0.632 (0.627-0.638) | 0.675 (0.668-0.683) | 0.649 (0.643-0.655) | 0.681 (0.669-0.693) | 0.651 (0.645-0.656) | 0.689 (0.680-0.697) | 0.653 (0.647-0.659) | 0.679 (0.668-0.689) | 0.663 (0.657-0.668) | 0.661 (0.655-0.666) | 0.664 (0.654-0.674) |
| Intercept & slope | 0.666 (0.661-0.671) | 0.665 (0.598-0.733) | 0.636 (0.624-0.648) | 0.632 (0.627-0.638) | 0.676 (0.668-0.683) | 0.649 (0.642-0.655) | 0.683 (0.671-0.695) | 0.651 (0.645-0.656) | 0.688 (0.680-0.696) | 0.654 (0.648-0.660) | 0.679 (0.668-0.689) | 0.663 (0.657-0.668) | 0.660 (0.655-0.666) | 0.664 (0.654-0.674) |
| eGFR | | | | | | | | | | | | | | |
| Mean & SD | 0.667 (0.662-0.672) | 0.653 (0.584-0.722) | 0.636 (0.624-0.648) | 0.635 (0.629-0.640) | 0.677 (0.669-0.684) | 0.651 (0.645-0.658) | 0.682 (0.670-0.694) | 0.653 (0.647-0.658) | 0.689 (0.681-0.698) | 0.655 (0.649-0.661) | 0.681 (0.671-0.692) | 0.664 (0.658-0.669) | 0.662 (0.657-0.668) | 0.665 (0.655-0.675) |
| Median & IQR | 0.667 (0.662-0.672) | 0.645 (0.577-0.713) | 0.635 (0.623-0.647) | 0.634 (0.629-0.640) | 0.676 (0.669-0.684) | 0.651 (0.644-0.657) | 0.682 (0.670-0.694) | 0.652 (0.647-0.658) | 0.689 (0.681-0.697) | 0.655 (0.649-0.661) | 0.681 (0.671-0.692) | 0.664 (0.658-0.669) | 0.662 (0.656-0.667) | 0.665 (0.655-0.675) |
| Intercept & slope | 0.665 (0.660-0.670) | 0.653 (0.587-0.718) | 0.634 (0.622-0.646) | 0.632 (0.626-0.638) | 0.675 (0.668-0.683) | 0.648 (0.642-0.655) | 0.682 (0.670-0.694) | 0.650 (0.645-0.656) | 0.688 (0.680-0.696) | 0.653 (0.647-0.659) | 0.679 (0.668-0.689) | 0.662 (0.657-0.668) | 0.660 (0.655-0.666) | 0.663 (0.653-0.673) |

Data are presented as C-index (95%CI), C-index change (95%CI) or NRI (95%CI) for MACE: Ischemic heart disease, stroke, heart failure or CVD-mortality.

Note: The trajectory measures were calculated including all recorded measurements within the 3 years prior to the baseline index date of 1st of January 2015. (2012, 2013 and 2014) and paired as mean & SD, median & IQR, and intercept & slope from a fitted growth model. The trajectory measure models were fitted using an offset procedure, sequentially adding one of the three types of paired trajectory measures to the respective (sub)sample reference model: mean & SD, median & IQR, and intercept & slope. All reference models (Cox-regression) included baseline measurements for age, age at T2D diagnosis, sex, HbA1c, LDL-cholesterol, HDL-cholesterol, eGFR, blood pressure lowering, lipid lowering, oral glucose lowering medication and insulin use.

HbA1c: Hemoglobulin A1c, LDL: low density lipoprotein, eGFR: estimated glomerular filtration rate, NRI: Net Reclassification Index, CI: Confidence Interval, SD: Standard deviation, IQR: Interquartile range.

Table S8. Performance improvement and reclassification statistics of trajectory measure updated prediction models, stratified by age group (<40/40-60/>60), sex and blood pressure lowering, lipid lowering, oral glucose lowering medication, and insulin use.

| Metric & Prediction model | Age | | | Sex | | Blood pressure lowering medication | | Lipid lowering medication | | Oral glucose lowering medication | | Insulin | |
|---|---|---|---|---|---|---|---|---|---|---|---|---|---|
| | <40 | 40-60 | >60 | Female | Male | No | Yes | No | Yes | No | Yes | No | Yes |
| **C-index change (95%CI)** | | | | | | | | | | | | | |
| HbA1c | | | | | | | | | | | | | |
| Mean & SD | 0.001 (-0.004-0.006) | 0.000 (-0.001-0.001) | 0.001 (0.000-0.002) | 0.001 (0.000-0.001) | 0.001 (0.000-0.001) | 0.000 (-0.001-0.001) | 0.001 (0.000-0.001) | 0.001 (0.000-0.001) | 0.001 (0.000-0.001) | 0.002 (0.001-0.003) | 0.001 (0.000-0.001) | 0.001 (0.000-0.001) | 0.002 (0.000-0.003) |
| Median & IQR | 0.000 (-0.007-0.008) | 0.001 (-0.000-0.002) | 0.001 (0.000-0.001) | 0.000 (-0.000-0.001) | 0.001 (0.000-0.002) | 0.000 (-0.001-0.001) | 0.001 (0.000-0.001) | 0.001 (-0.000-0.001) | 0.001 (0.000-0.001) | 0.001 (0.000-0.002) | 0.001 (0.000-0.001) | 0.001 (0.000-0.001) | 0.002 (0.000-0.003) |
| Intercept & slope | 0.000 (-0.005-0.007) | 0.001 (-0.001-0.002) | 0.001 (0.000-0.001) | 0.001 (-0.000-0.001) | 0.001 (0.000-0.002) | 0.001 (-0.000-0.003) | 0.001 (0.000-0.001) | 0.001 (-0.000-0.002) | 0.001 (0.000-0.001) | 0.000 (-0.000-0.001) | 0.001 (0.000-0.001) | 0.001 (0.000-0.001) | 0.001 (-0.000-0.003) |
| LDL-cholesterol | | | | | | | | | | | | | |
| Mean & SD | 0.022 (0.002-0.042) | 0.002 (-0.000-0.004) | 0.000 (0.000-0.001) | 0.001 (0.000-0.001) | 0.001 (0.000-0.002) | 0.000 (-0.000-0.001) | 0.001 (0.000-0.001) | 0.001 (0.000-0.001) | 0.001 (0.000-0.001) | 0.001 (0.000-0.002) | 0.001 (0.000-0.001) | 0.001 (0.000-0.001) | 0.001 (-0.000-0.001) |
| Median & IQR | 0.038 (0.003-0.073) | 0.002 (-0.001-0.003) | 0.000 (-0.000-0.001) | 0.001 (-0.000-0.001) | 0.001 (0.000-0.001) | 0.000 (-0.000-0.001) | 0.001 (0.000-0.001) | 0.001 (0.000-0.001) | 0.001 (0.000-0.001) | 0.000 (-0.000-0.001) | 0.001 (-0.000-0.001) | 0.001 (0.000-0.001) | 0.001 (-0.000-0.001) |
| Intercept & slope | 0.017 (0.000-0.033) | 0.001 (-0.000-0.003) | 0.000 (-0.000-0.001) | 0.001 (0.000-0.001) | 0.001 (0.000-0.001) | 0.002 (-0.000-0.003) | 0.000 (0.000-0.001) | 0.000 (-0.000-0.000) | 0.001 (0.000-0.002) | 0.000 (-0.000-0.001) | 0.001 (0.000-0.001) | 0.001 (0.000-0.001) | 0.001 (-0.000-0.002) |
| eGFR | | | | | | | | | | | | | |
| Mean & SD | 0.005 (-0.019-0.037) | 0.001 (-0.001-0.004) | 0.003 (0.002-0.004) | 0.002 (0.001-0.003) | 0.003 (0.001-0.004) | 0.001 (-0.000-0.002) | 0.002 (0.002-0.003) | 0.001 (0.001-0.002) | 0.003 (0.001-0.004) | 0.003 (0.001-0.005) | 0.002 (0.001-0.003) | 0.002 (0.001-0.003) | 0.002 (0.000-0.003) |
| Median & IQR | -0.003 (-0.015-0.011) | 0.001 (-0.001-0.002) | 0.002 (0.001-0.004) | 0.001 (0.000-0.002) | 0.003 (0.001-0.004) | 0.001 (-0.000-0.002) | 0.002 (0.001-0.003) | 0.001 (0.000-0.001) | 0.003 (0.002-0.004) | 0.003 (0.001-0.005) | 0.002 (0.001-0.002) | 0.002 (0.001-0.003) | 0.001 (0.000-0.003) |
| Intercept & slope | 0.004 (-0.007-0.015) | 0.000 (-0.001-0.001) | 0.000 (-0.000-0.001) | 0.000 (-0.000-0.001) | 0.000 (-0.000-0.001) | 0.001 (-0.000-0.002) | 0.000 (-0.000-0.000) | 0.000 (-0.000-0.000) | 0.000 (-0.000-0.001) | 0.001 (-0.000-0.002) | 0.000 (-0.000-0.000) | 0.000 (-0.000-0.001) | 0.000 (-0.000-0.001) |
| **Continuous NRI (95%CI)** | | | | | | | | | | | | | |
| HbA1c | | | | | | | | | | | | | |
| Mean & SD | 13.7 (- | 3.9 (- | 8.0 (4.7- | 5.7 (-0.7- | 4.3 (0.6- | 6.2 (- | 5.8 (2.9- | 3.2 (-2.2- | 5.9 (2.2- | 10.2 | 4.4 (0.9- | 5.8 (1.8- | 5.8 (-4.9- |

| | | | | | | | | | | | | | |
|---|---|---|---|---|---|---|---|---|---|---|---|---|---|
| | 30.9-40.6) | 6.4-10.4) | 12.9) | 10.8) | 8.2) | 2.6-14.1) | 10.4) | 8.6) | 9.6) | (1.7-18.0) | 9.3) | 10.3) | 11.4) |
| Median & IQR | -1.3 (-40.6-42.5) | 6.5 (-2.5-15.0) | 7.0 (3.4-10.9) | 3.8 (-2.6-8.7) | 5.0 (1.7-8.6) | 5.6 (-4.0-13.1) | 5.4 (1.3-9.7) | 2.7 (-3.7-10.4) | 6.1 (2.2-10.6) | 6.6 (-5.8-15.0) | 5.0 (0.5-9.8) | 5.1 (1.6-8.7) | 9.6 (-1.5-15.1) |
| Intercept & slope | 7.5 (-27.5-40.1) | 7.5 (0.3-15.0) | 8.6 (4.0-13.4) | 6.2 (-0.8-11.1) | 7.5 (2.5-11.1) | 9.8 (-1.1-17.1) | 7.0 (4.0-10.9) | 7.7 (-1.7-13.6) | 8.3 (2.7-12.5) | 6.7 (-2.8-15.4) | 7.2 (3.3-10.8) | 8.9 (4.9-12.6) | 8.8 (-0.9-14.9) |
| LDL-cholesterol | | | | | | | | | | | | | |
| Mean & SD | 23.6 (-36.7-73.7) | 9.7 (2.2-17.4) | 4.3 (0.5-7.5) | 3.2 (-3.2-8.7) | 4.3 (-0.5-8.3) | 5.9 (-3.0-14.9) | 4.5 (1.8-8.3) | 1.8 (-5.3-7.7) | 2.8 (-0.7-7.5) | 4.8 (-3.6-11.2) | 3.5 (-0.6-7.4) | 4.5 (0.7-7.9) | 1.5 (-5.9-7.8) |
| Median & IQR | 23.9 (-20.2-57.5) | 8.1 (1.9-17.3) | 3.5 (-0.1-7.0) | 2.6 (-4.8-8.3) | 3.8 (-1.1-6.8) | 7.0 (-1.2-14.5) | 3.9 (0.3-8.5) | 1.9 (-6.2-10.0) | 1.6 (-2.4-6.7) | 3.9 (-4.8-9.6) | 3.0 (-0.6-6.5) | 4.6 (-0.1-8.3) | -1.6 (-10.2-7.2) |
| Intercept & slope | 34.1 (-2.3-64.9) | 6.7 (-2.6-18.3) | 6.3 (1.9-10.8) | 6.7 (-0.9-13.4) | 5.9 (0.8-9.4) | 12.9 (4.6-20.6) | 4.5 (1.3-9.0) | 3.9 (-4.6-10.4) | 6.5 (1.9-10.9) | 6.0 (-1.6-14.3) | 5.6 (1.6-9.9) | 5.4 (1.6-10.3) | 6.8 (-5.0-15.3) |
| eGFR | | | | | | | | | | | | | |
| Mean & SD | 13.7 (-25.2-45.5) | 8.4 (-1.7-14.7) | 7.3 (2.5-11.3) | 8.5 (3.1-14.4) | 7.2 (2.3-11.7) | 6.2 (-2.3-15.2) | 7.3 (3.5-11.1) | 4.7 (-0.4-11.9) | 8.0 (4.0-12.0) | 7.7 (-0.2-15.2) | 6.9 (3.0-11.5) | 8.4 (5.4-12.2) | 5.3 (-2.2-14.4) |
| Median & IQR | -2.2 (-35.2-47.2) | 2.9 (-5.6-10.8) | 7.1 (2.8-11.9) | 8.2 (2.6-13.8) | 8.0 (3.8-12.6) | 7.5 (-2.2-16.3) | 7.2 (3.2-11.8) | 4.0 (-1.8-11.5) | 8.9 (4.9-13.5) | 8.9 (-0.7-19.5) | 6.9 (3.3-10.3) | 7.9 (5.0-11.8) | 8.3 (0.8-16.7) |
| Intercept & slope | 12.2 (-40.0-47.8) | -7.2 (-17.1-0.4) | 3.0 (-2.6-8.3) | 3.6 (-6.4-10.7) | 4.6 (-3.7-10.3) | 9.5 (-3.8-20.5) | 2.8 (-2.3-7.5) | 8.5 (-2.6-18.1) | 2.2 (-3.2-9.4) | 3.7 (-6.6-13.3) | 2.8 (-3.0-7.8) | 6.6 (0.3-11.9) | 1.8 (-5.6-12.4) |

Data are presented as C-index (95%CI), C-index change (95%CI) or NRI (95%CI) for MACE: Ischemic heart disease, stroke, heart failure or CVD-mortality.
Note: The trajectory measures were calculated including all recorded measurements within the 3 years prior to the baseline index date of 1st of January 2015. (2012, 2013 and 2014) and paired as mean & SD, median & IQR, and intercept & slope from a fitted growth model. The trajectory measure models were fitted using an offset procedure, sequentially adding one of the three types of paired trajectory measures to the respective (sub)sample reference model: mean & SD, median & IQR, and intercept & slope. All reference models (Cox-regression) included baseline measurements for age, age at T2D diagnosis, sex, HbA1c, LDL-cholesterol, HDL-cholesterol, eGFR, blood pressure lowering, lipid lowering, oral glucose lowering medication and insulin use.
HbA1c: Hemoglobulin A1c, LDL: low density lipoprotein, eGFR: estimated glomerular filtration rate, NRI: Net Reclassification Index, CI: Confidence Interval, SD: Standard deviation, IQR: Interquartile range.

Figure S1. Cumulative incidence curves of MACE endpoint for total study sample, and stratified by sex, age group and blood pressure lowering, lipid lowering, oral glucose lowering medication, and insulin use.

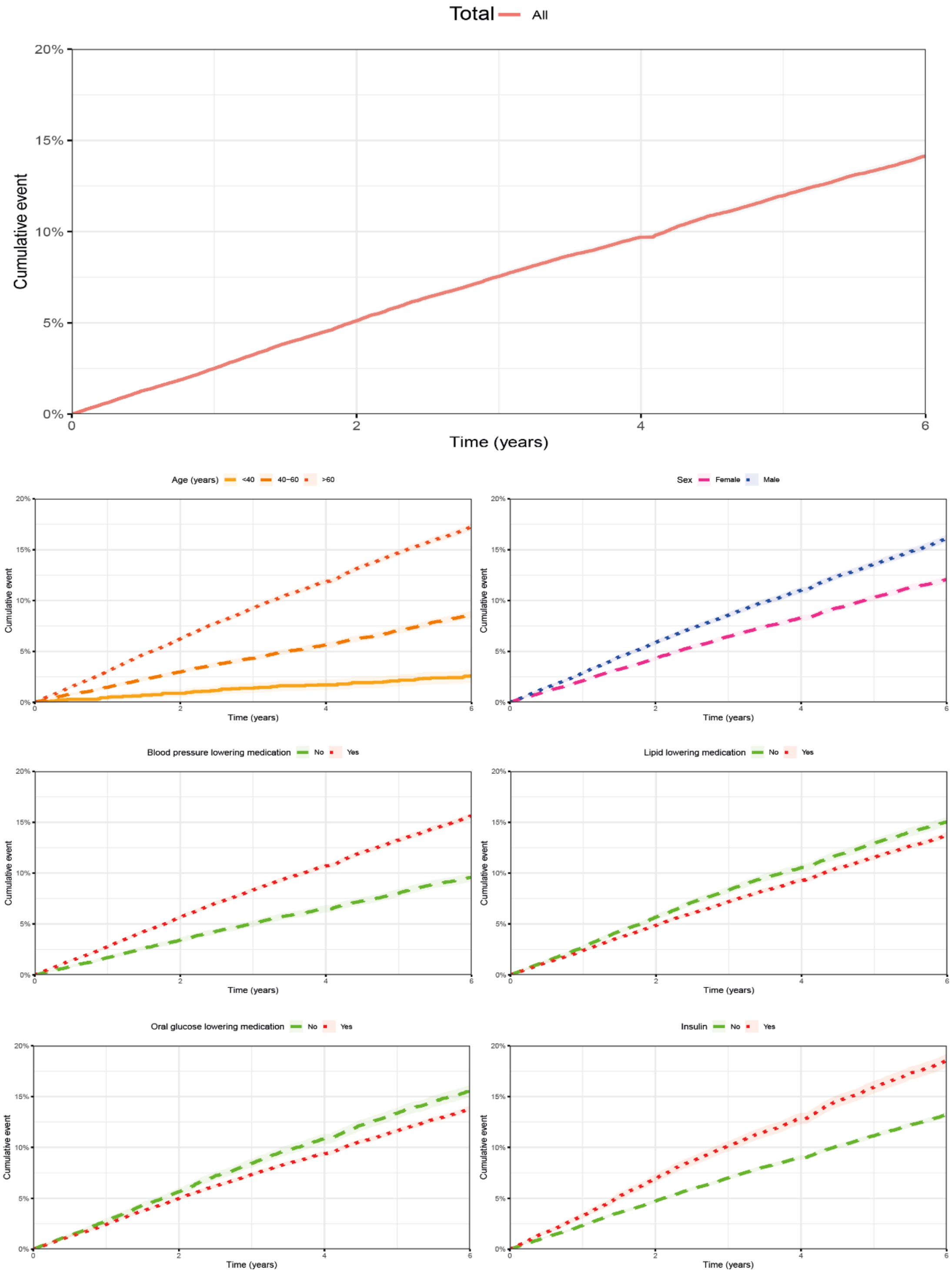
